# The Role of Network Connectivity and Transcriptomic Vulnerability in Shaping Grey Matter Atrophy in Multiple Sclerosis

**DOI:** 10.64898/2026.02.13.26346243

**Authors:** Mar Barrantes-Cepas, Mario Tranfa, David R. van Nederpelt, Ismail Koubiyr, Luigi Lorenzini, Birgit Helmlinger, Stefan Ropele, Daniela Pinter, Christian Enzinger, Tomas Uher, Manuela Vaneckova, Joep Killestein, Eva M.M. Strijbis, Martijn D. Steenwijk, Hugo Vrenken, Frederik Barkhof, Menno M. Schoonheim, Giuseppe Pontillo

**Author notes:** Corresponding author: Mar Barrantes-Cepas. These authors contributed equally to this work (Menno M. Schoonheim, Giuseppe Pontillo).

## Abstract

Clinical progression is strongly linked to grey matter atrophy in multiple sclerosis (MS), detectable early on MRI and progressing non-randomly across the brain. However, the mechanisms driving its spatio-temporal progression and individual variability remain unclear. Using MRIs from 2,187 participants, alongside normative data, we systematically investigated network-based mechanisms underlying MS-related atrophy. Regional atrophy colocalised with functional cortical hubs, supporting the *nodal stress* hypothesis, and propagated along anatomical and functional connections, consistent with *transneuronal degeneration*. *Lesional disconnection* and *transcriptomic vulnerability* played marginal roles. Patient- and subgroup-level analyses revealed that network-based mechanisms are specifically linked to MS-related neurodegeneration and may operate differently in distinct subtypes or disease phases. Atrophy patterns were anchored to the connectivity profiles of disease epicentres involving the visual, sensorimotor, and temporal cortices, and the hippocampi and thalami. Network-based measures enhanced the prediction of future atrophy progression in individual with MS, providing a mechanistic framework to understand neurodegeneration in MS.

## Introduction

Clinical progression in multiple sclerosis (MS) is strongly related to grey matter (GM) atrophy, considered as a marker of neurodegeneration and correlating with both physical disability and cognitive impairment (*1–3*). The spatiotemporal evolution of regional GM atrophy in MS has been extensively explored: brain tissue volume loss is present since the very early phases of the disease, progresses with time, and does not occur uniformly throughout the brain, with regions such as the thalamus and the cingulate cortex shrinking faster and earlier than others (*4*). Additionally, there is substantial inter-subject heterogeneity in MS-related atrophy patterns, which seem to cluster around distinct “modes” of neurodegeneration that do not necessarily align with conventional clinical phenotypes (*5*).

While current evidence clearly highlights that GM atrophy in MS is largely non-random (*6*), the mechanisms that drive these spatial and temporal patterns of neurodegeneration remain poorly understood, particularly from an individual-level perspective (*1, 2, 7–9*), hindering the development of targeted therapeutic strategies aimed at slowing the neurodegenerative component of the disease (*10*). Indeed, previous studies have identified where and when MS-related neurodegeneration occurs, focusing primarily on regional characterization and group-level comparisons (*4, 6, 9, 11*). However, they have only provided limited understanding of the processes underlying GM atrophy and why certain people with MS (pwMS) are more susceptible than others.

Moving beyond regional analyses, a network-based perspective enables the shift in focus to the broader organisational principles of the brain that may underlie vulnerability to disease (*12–14*). Recent advances in network neuroscience and transcriptomics, along with the increasing availability of public data resources in these domains, have facilitated the development of a more integrative approach that can now elucidate mechanisms underlying neurodegeneration based on structural and functional connectivity, as well as gene expression profiles from normative data (*15–18*). Leveraging this network-based approach, evidence across neurologic and psychiatric disorders suggests that neurodegeneration follows network-constrained patterns (*17–19*).

Key mechanisms hypothesised to explain spatiotemporal atrophy patterns include *nodal stress*, *transneuronal degeneration*, *transcriptomic vulnerability*, and *lesional disconnection* (*1, 2, 7, 8, 14, 20, 21*). Specifically, highly connected regions (i.e., hubs) may be more prone to neurodegeneration, due to their greater metabolic demand leading to cellular dysfunction, a concept known as *nodal stress* (*1*). Additionally, atrophy often mirrors the brain’s intrinsic connectivity, suggesting that damage may spread to functionally and structurally connected regions (i.e., neighbours) via *transneuronal degeneration* (*3*). Finally, this network-based degeneration might be specifically driven by axonal transection through focal lesions, leading to secondary, tract-mediated degeneration of connected GM regions (*lesional disconnection*) (*14, 22*). Complementary to these connectivity-based explanations, the concept of *transcriptomic vulnerability* entails that regions with similar gene expression profiles might have similar biological properties, hence tending to be similarly affected by disease processes (*23–25*).

These hypotheses have also been proposed and investigated to some extent in MS (*8*). In particular, GM atrophy has been often correlated with WM disruption (*10, 20*). Unfortunately, these mechanisms have been studied in isolation, using small, single-site cohorts, with varying imaging modalities and/or analytic methods (*4, 6, 9, 21*). Consequently, their relative contributions to MS-related GM atrophy remain unclear.

Here, we used an integrated approach to systematically investigate the above-mentioned mechanisms in a large multicentric dataset. First, we explored these mechanisms at the group level. Second, we performed subgroup- and individual-level analyses to capture interindividual variability and to determine whether certain patient subgroups may be more susceptible to specific mechanisms. Finally, we investigated the potential of such a network-based approach to predict future atrophy progression.

## Results

A schematic overview of the study workflow is shown in Fig. 1.

**Fig. 1.**
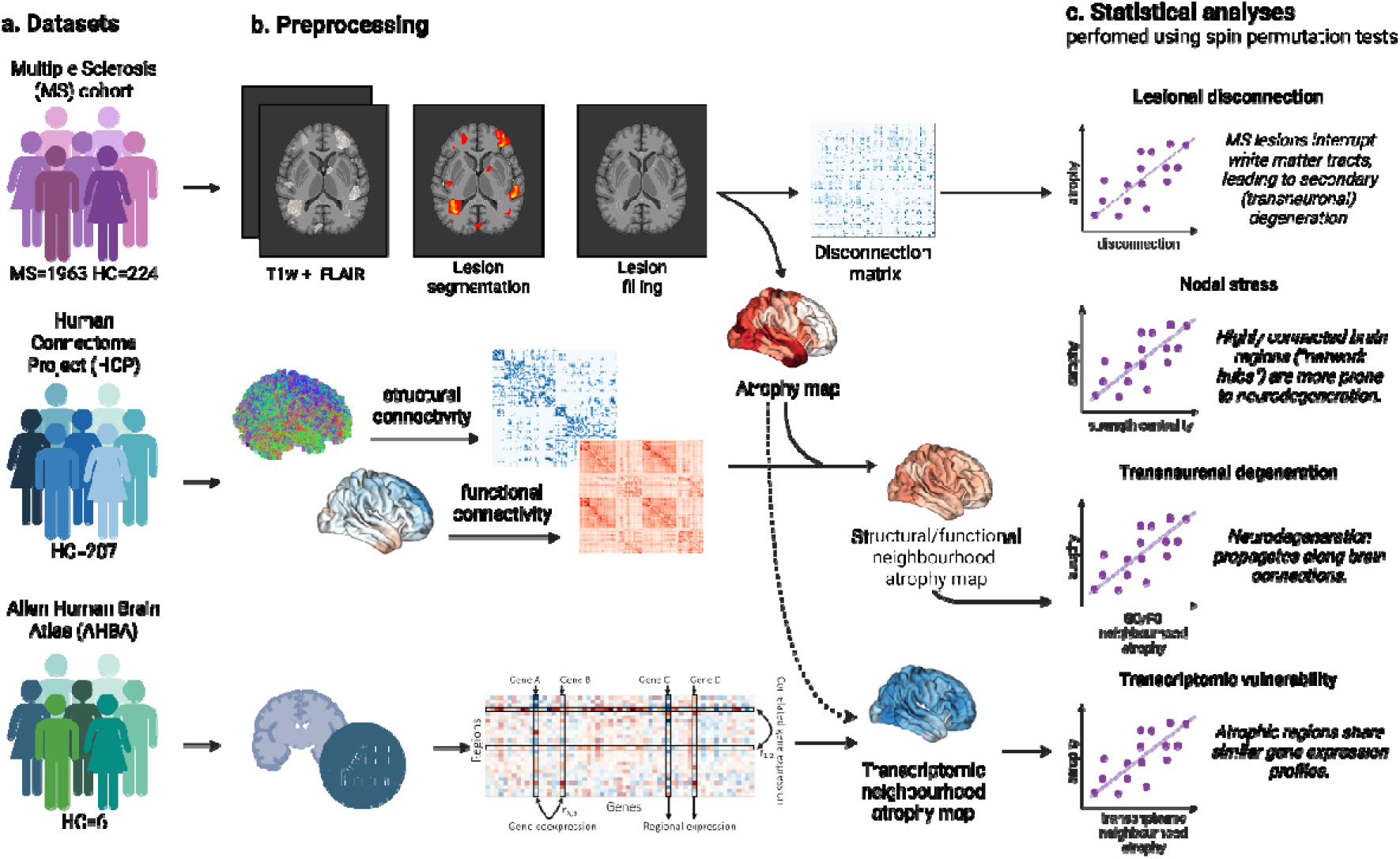
Study workflow. (a) The study integrated three datasets: MS Cohort and healthy controls, including T1-weighted (T1w) and FLAIR scans; the Human Connectome Project (HCP), providing structural and functional connectomes (SC/FC); and the Allen Human Brain Atlas (AHBA), providing gene expression data. (b) In the MS cohort, lesions were segmented using LST-AI and subsequently T1w scans were filled with NiftySeg. Disconnectome matrices were computed using the Lesion Quantification Toolkit (LQT), while cortical thickness and subcortical volumes were extracted with FreeSurfer. Atrophy maps were generated using Cohen’s *d*, and connectome and gene expression data were preprocessed using the ENIGMA toolbox. Neighbouring atrophy maps were obtained as described in Methods (*18*). (c) Statistical analyses included spin permutation tests (n=10,000) to assess spatial correspondence. Created using https://BioRender.com and adapted from (*15, 16, 18*).

### Study population

We retrospectively collected and analysed structural brain MRIs and clinico-demographic data of pwMS or clinically isolated syndrome (CIS), as well as of healthy controls (HCs), from three European centres, acquired between 2014 and 2023. The final study population consisted of 2,187 participants (1,963 pwMS/CIS, 70% female, 45.3±9.8 years; 224 HCs, 62.5% female, 41.1±12.4 years). For the 565 pwMS/CIS for whom more than one MRI scan was available (total number of sessions = 1245, mean latest follow-up duration = 3.48 years, range: 0.5 – 8.8 years), the first scan was used to model cross-sectional atrophy, while all timepoints were used to model longitudinal atrophy. Demographic and clinical characteristics of the studied population are given in Table 1, while the different acquisition protocols are detailed in Table S1 and further information regarding image quality control and excluded participants is provided in Fig. S1 and Table S2.

**Table 1.** Demographic, clinical and MRI characteristics of the studied population at baseline.

| | HC | pwMS/CIS | $p$-value |
| --- | --- | --- | --- |
| N | 224 | 1963 |  |
| Age [years], mean±SD | 41.1±12.4 | 45.3±9.84 | < 0.001* |
| Female, n (%) | 140 (62.5%) | 1375 (70%) | 0.02<sup>†</sup> |
| **Disease characteristics** |  |  |  |
| Phenotype, n (%) |  |  |  |
| CIS |  | 57 (2.9%) | - |
| RRMS |  | 1620 (82.5%) | - |
| SPMS |  | 242 (12.3%) | - |
| PPMS |  | 44 (2.2%) | - |
| Disease duration [years], mean±SD |  | 14.5±9.2 | - |
| EDSS, median (range) |  | 2.5 [0 – 8.5] | - |
| DMT, n (%): yes/no |  | 1505 (76.7%) / 458 (23.3%) | - |
| TLV [mL], median (IQR) |  | 3.36 [1.07 – 9.24] | - |
| BPF, mean±SD | 0.77±0.02 | 0.75±0.05 | < 0.001* |

Abbreviations: BPF = brain parenchymal fraction, CIS = clinically isolated syndrome, DMT = disease-modifying treatment, EDSS = Expanded Disability Status Scale, HC = healthy controls, IQR = interquartile range, PPMS = primary-progressive multiple sclerosis, pwMS/CIS = people with multiple sclerosis or clinically isolated syndrome, RRMS = relapsing-remitting multiple sclerosis, SD = standard deviation, SPMS = secondary-progressive multiple sclerosis, TLV = total lesion volume.
\*Permutation t-test (n = 10,000); †Chi-square test.

### Grey matter atrophy in MS

We processed 3D T1-weighted (T1w) and 3D Fluid-Attenuated Inversion Recovery (FLAIR) scans to obtain values of thickness from 100 cortical regions (Schaefer atlas (*26*)) and volume from 14 subcortical structures (FreeSurfer’s Aseg atlas (*27*)). Individual regional atrophy was expressed as W-scores, reflecting the deviation from the HCs group while adjusting for the physiological effects of age, sex, intracranial volume (for subcortical volumes), and site. When comparing W-score maps (sign-flipped so that more positive scores indicated greater atrophy), pwMS/CIS exhibited widespread reduction of subcortical GM volumes relative to HCs, most pronounced in the thalami (Cohen’s *d* = 1.07, *p*_FDR_ < 0.001). Significant cortical thinning was evident in the temporo-polar, sensorimotor, posterior cingulate, and precuneal cortices (Fig. 2).

**Fig. 2.**
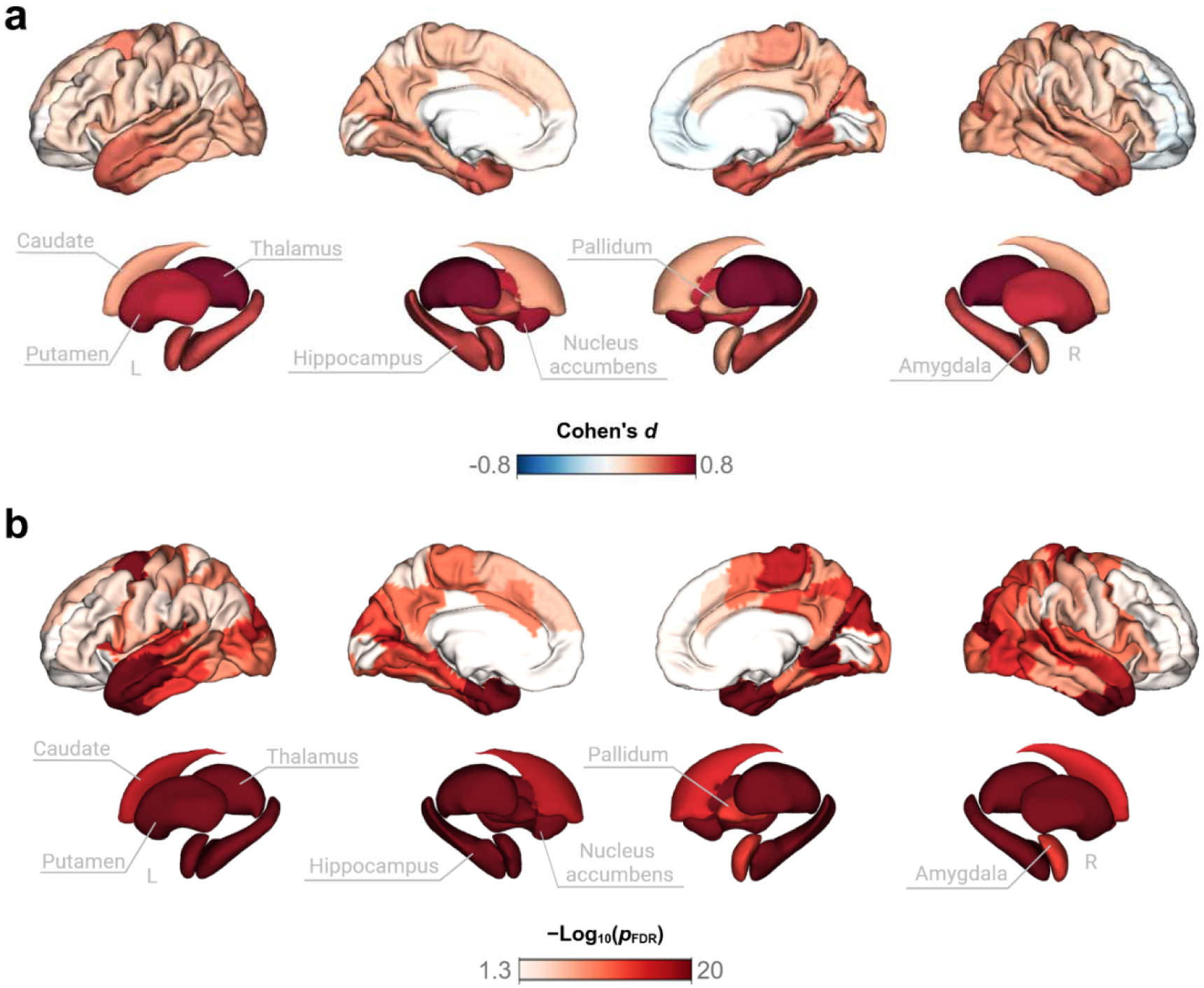
Grey matter atrophy in pwMS/CIS. (a) Atrophy maps show cross-sectional differences (Cohen’s *d*) in cortical thickness and subcortical volumes between pwMS/CIS and HCs, adjusting for physiological effects of age, sex, intracranial volume (for subcortical volumes), and site. (b) Negative log10-transformed FDR-corrected *p-*values are shown.

### Network mechanisms shape grey matter atrophy

We operationalised the *nodal stress*, *transneuronal degeneration*, *lesional disconnection*, and *transcriptomic vulnerability* hypotheses in terms of spatial similarity between regional atrophy and a set of network-based maps. Statistical significance of spatial correlations was assessed against null distributions obtained by spatially permuting maps randomly (10,000 repetitions), with significance set at *p* < 0.05.

To investigate the *nodal stress* hypothesis (i.e., highly connected regions are more prone to neurodegeneration), we investigated the association between regional GM atrophy and functional and structural hubness maps, obtained using normative data derived from 207 healthy adults sampled from the Human Connectome Project (HCP), and expressed in terms of strength centrality (i.e., the sum of all cortico-cortical or subcortico-cortical edges insisting on each node)(Fig. 3a, b). We found a positive correlation between functional cortico-cortical strength centrality and cortical atrophy (*r* = 0.38, *p* = 0.01; Fig. 3c), indicating that cortical regions with stronger functional hub characteristics tended to exhibit greater atrophy. In contrast, no associations with regional atrophy were found for functional subcortico-cortical strength centrality (*r* = 0.31, *p* = 0.15; Fig. 3c), structural cortico-cortical strength centrality (*r* = –0.15, *p* = 0.21; Fig. 3d), or structural subcortico-cortical strength centrality (*r* = 0.35, *p* = 0.11; Fig. 3d).

**Fig. 3.**
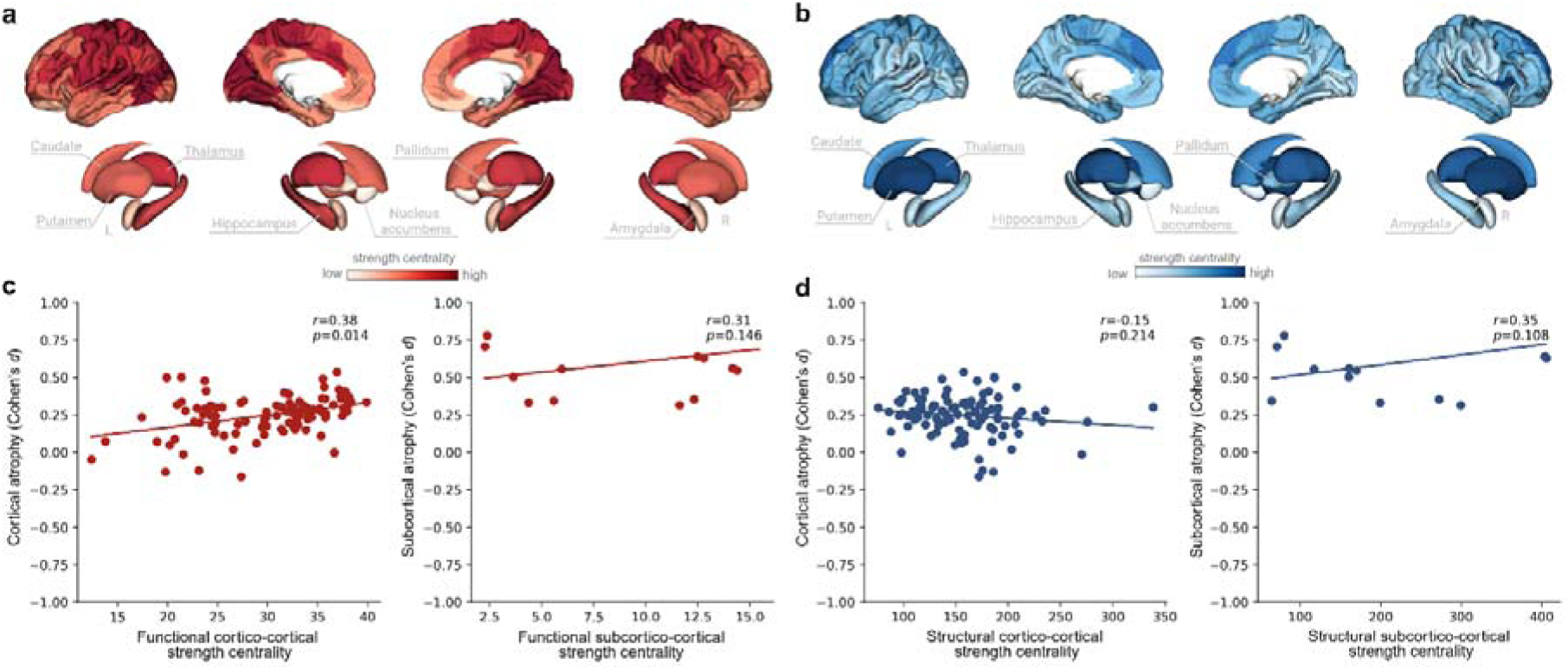
Functional cortical hubs exhibit greater GM atrophy in pwMS/CIS. (a) Functional and (b) structural strength centrality maps obtained using the HCP data. (c) Spatial correlation between functional and (d) structural strength centrality and grey matter atrophy.

For *transneuronal degeneration* (i.e., neurodegeneration propagates along brain connections), we tested the association between regional GM atrophy and neighbourhood atrophy maps, defined as the average atrophy of each region’s first-degree neighbours, weighted by the strength of their functional or structural connections (Fig. 4a, b) (*18*). We found a positive correlation between cortical atrophy and both functional (*r* = 0.41, *p* = 0.005; Fig. 4c) and structural (*r* = 0.51, *p* = 0.004; Fig. 4d) cortico-cortical neighbourhood atrophy, indicating that cortical regions tend to lose volume together with their functionally- and structurally-connected neighbours. No associations with atrophy were observed for subcortical regions and functional (*r* = 0.29, *p* = 0.16; Fig. 4c) or structural (*r* = 0.13, *p* = 0.35; Fig. 4d) neighbourhood atrophy.

**Fig. 4.**
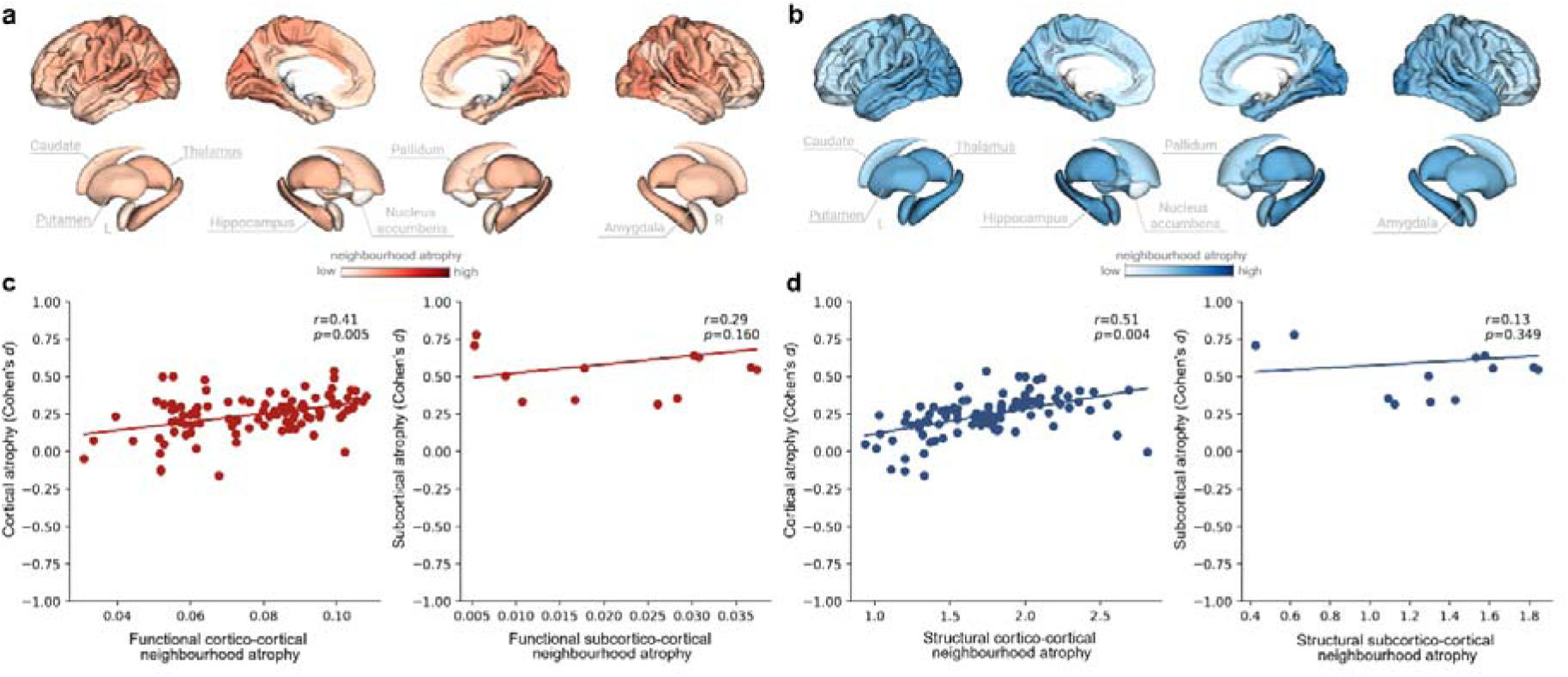
Connected cortical regions tend to atrophy together in pwMS/CIS. (a) Functional and (b) structural neighbourhood atrophy maps, defined as the average atrophy of each region’s first-degree neighbours, weighted by connection strength. (c) Spatial correlation between functional and (d) structural neighbourhood atrophy and grey matter atrophy.

We then investigated whether atrophy overlapped with *lesional disconnection* (i.e., WM lesions drive secondary neurodegeneration in anatomically connected regions). First, we obtained data-driven structural disconnectomes from lesion masks and the HCP1065 tractography atlas using the Lesion Quantification Toolkit (LQT) (*28*). Then, we tested the spatial correlation between regional GM atrophy and node disconnection, expressed as the regional strength centrality of the disconnectome matrix (i.e., the sum of all cortico-cortical or subcortico-cortical edges insisting on each node) (Fig. 5a). On average, lesion-related disconnection was more prominent in the deep GM and in temporal and posterior cortical regions (Fig. 5a). We found no relationship between cortical GM atrophy and structural disconnection (*r* = 0.16, *p* = 0.1; Fig. 5c). Similarly, no relationship was observed between subcortical atrophy and structural disconnection (*r* = 0.10, *p* = 0.35; Fig. 5c).

**Fig. 5.**
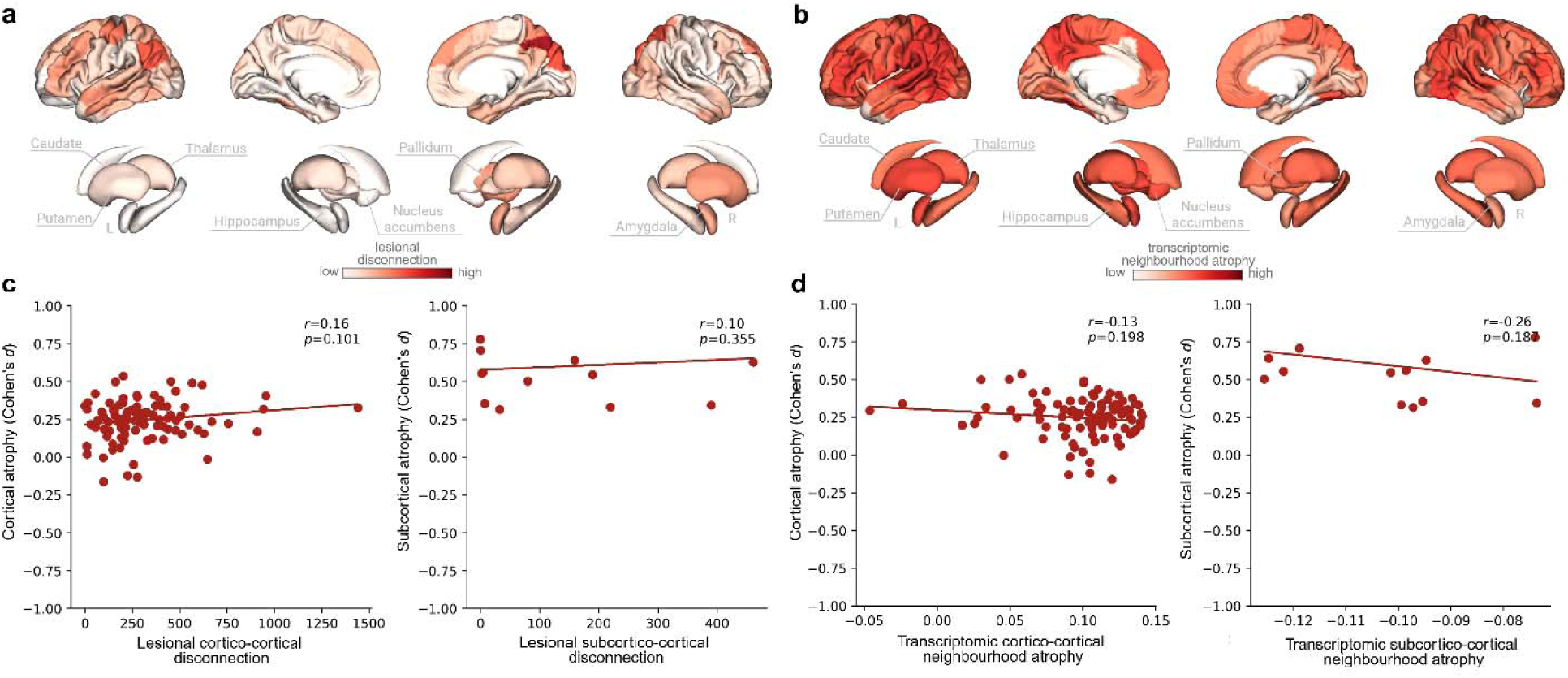
No associations between GM atrophy and lesional disconnection or transcriptomic vulnerability in pwMS/CIS. (a) MS lesion disconnection maps obtained with a normative tractography atlas. (b) Transcriptomic neighbourhood atrophy, defined as the average atrophy of each region’s first-degree neighbours, weighted by transcriptomic similarity. (c) Correlation of lesion-related disconnection and (d) transcriptomic neighbourhood atrophy and grey matter atrophy.

Finally, for *transcriptomic vulnerability* (i.e., regional neurodegeneration is mediated by shared gene expression profiles), we tested the association between regional GM atrophy and transcriptomic neighbourhood atrophy maps. The latter was defined as the average atrophy of each region’s first-degree neighbours, weighted by their normative transcriptomic similarity, derived from the Allen Human Brain Atlas (AHBA) (Fig. 5b). In the cortex, no relationship was found (*r* = –0.13, *p* = 0.2; Fig. 5d), as well as in subcortical regions (*r* = −0.26, *p* = 0.19; Fig. 5d).

### Patient- and subgroup-tailored atrophy modelling

We adapted our network-based models to assess whether they configured atrophy patterns in individual pwMS/CIS and specific subgroups. Specifically, we assessed the spatial correlations between individual atrophy (i.e., W-score) maps and network-based maps on a per-patient basis, and Fisher’s *z*-transformed the obtained *r* values to enable comparisons across individuals and subgroups.

At the individual level, all observed associations were significantly different from zero (*p*_FDR_ < 0.05 at one-sample *t*-tests), indicating relatively consistent effects across pwMS/CIS (Fig. 6a). The greatest effects were observed for the association between regional GM atrophy and both structural (Cohen’s *d* = 1.76) and functional (*d* = 0.61) cortico-cortical neighbourhood atrophy, supporting the role of *transneuronal degeneration* as the main mechanism shaping MS-related atrophy patterns. Associations between subcortical atrophy and network-based maps exhibited the greatest variability across pwMS/CIS, suggesting higher heterogeneity in the mechanisms driving deep GM neurodegeneration.

**Fig. 6.**
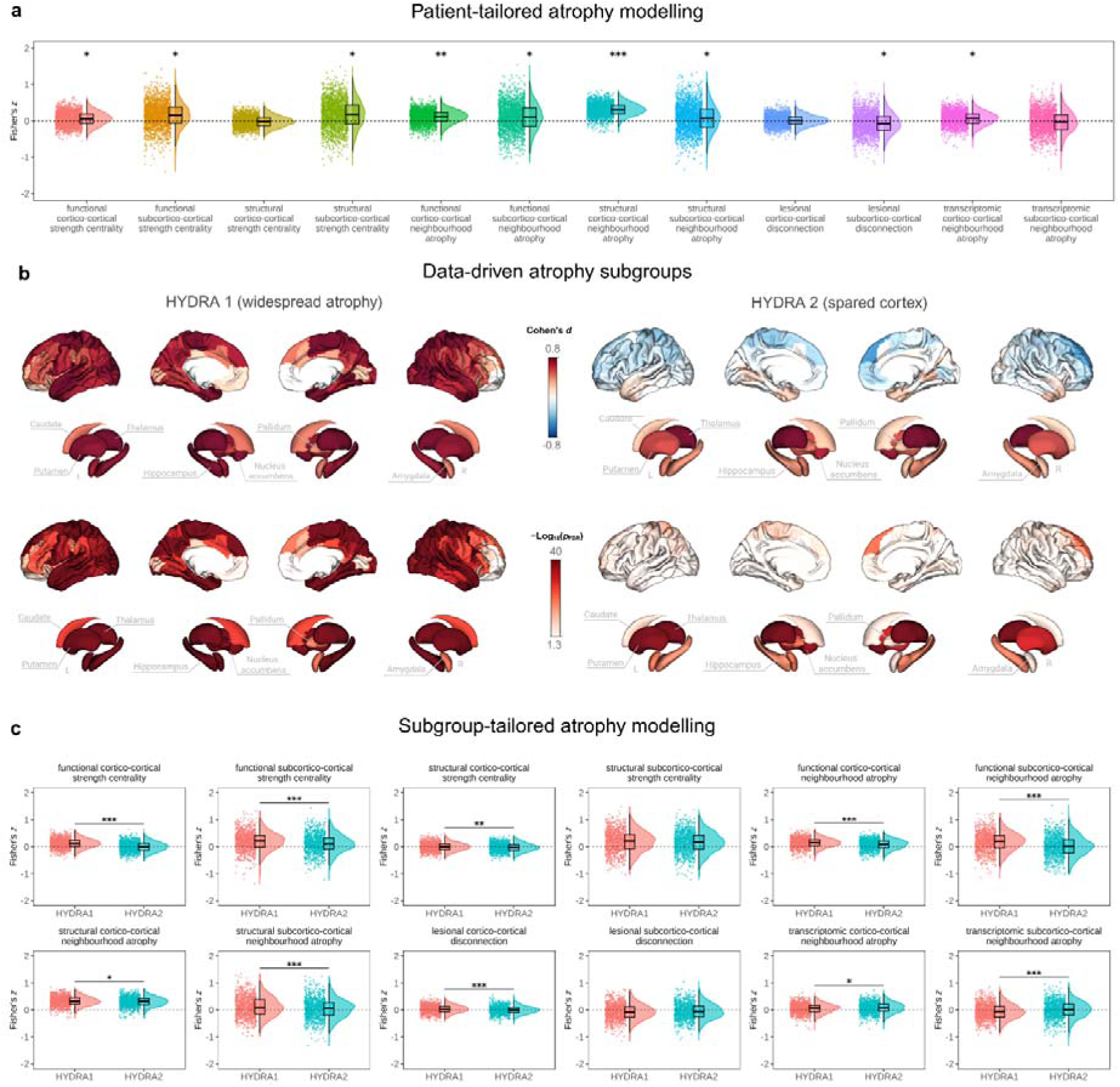
Patient- and subgroup-tailored atrophy modelling. (a) Fisher *z*-transformed individual correlation coefficients are shown for each mechanism. All mechanisms were significant against the null using one sample T-test and FDR-corrected *p*-values. Effect sizes are indicated as: * = small Cohen’s *d* (0.2 – 0.5), ** = moderate Cohen’s *d* (0.5 – 0.8), *** = large Cohen’s *d* (≥0.8). (b) Atrophy maps showing effect sizes (Cohen’s d) and negative log10-transformed FDR-corrected *p-*values for the data-driven HYDRA groups. (c) Boxplots comparing HYDRA subgroups across different mechanisms. *p* < 0.05 (*)*, p* < 0.01 (**)*, p* < 0.001 (***).

We then investigated the role of the different network-based mechanisms in shaping GM atrophy in distinct pwMS/CIS subgroups. First, we used the HYDRA algorithm, a non-linear semi-supervised machine learning approach, to subgroup patients into distinct regional atrophy profiles (*29*). We identified two main GM atrophy phenotypes: the first subgroup exhibited widespread cortical and subcortical atrophy, while the second showed selective subcortical GM volume loss, with relatively preserved cortical thickness (Fig. 6b and Fig. S2). PwMS/CIS assigned to the two clusters were slightly different in terms age (*p* < 0.001), sex (*p* < 0.001), lesion volume (*p* < 0.001), and clinical phenotype (*p* = 0.016), but did not differ in disease duration (*p* = 0.69) or physical disability level (*p* = 0.19) (Table S3). When investigating neurodegeneration mechanisms in the two data-driven subgroups, we observed that network-based mechanisms were consistently more relevant in the widespread atrophy subgroup than in individuals with spared cortical GM (Fig. 6c). Similarly, we compared neurodegeneration mechanisms across clinical phenotypes, defined as early relapsing (ER: CIS + relapsing-remitting MS (RRMS) with a disease duration below 5 years, N = 360), late relapsing (LR: RRMS with a disease duration above or equal to 5 years, N = 1317), secondary progressive (SP, N = 242) and primary progressive (PP, N = 44) (*30*). We observed that associations between GM atrophy and network-based maps (especially those indexing *nodal stress* and *transneuronal degeneration*) were stronger in more advanced (LR and SP) compared to earlier (ER) phases of the disease, with PP patients showing intermediate behaviour (Fig. S3).

### Data-driven disease epicentres

We investigated whether MS-related atrophy patterns were anchored to the connectivity profiles of specific regions. To identify these connectivity-based disease “epicentres” (i.e., critical regions where the disease process supposedly begins to then spread “transneuronally” to other brain regions facilitated by the underlying connectivity patterns), we used a previously adopted approach (*31*). Briefly, we correlated the structural and functional connectivity profiles of each brain region with individual atrophy maps and repeated this systematically across all regions. The resulting correlation coefficients were *r*-to-*z* transformed, and one-sample, one-tailed *t*-tests were performed for each region to assess their significance at the group level. Disease epicentres thus represented regions whose functional and structural connectivity profile spatially resembled MS-related atrophy maps.

Both structural and functional analyses revealed overlapping candidate epicentre regions within the visual and temporal cortices (Fig. 7). Cortical functional epicentres also extended to the sensorimotor cortex. Within subcortical structures, the thalamus, hippocampus, and amygdala were identified as structural epicentres of neurodegeneration, with the latter two also emerging as functional epicentres.

**Fig. 7.**
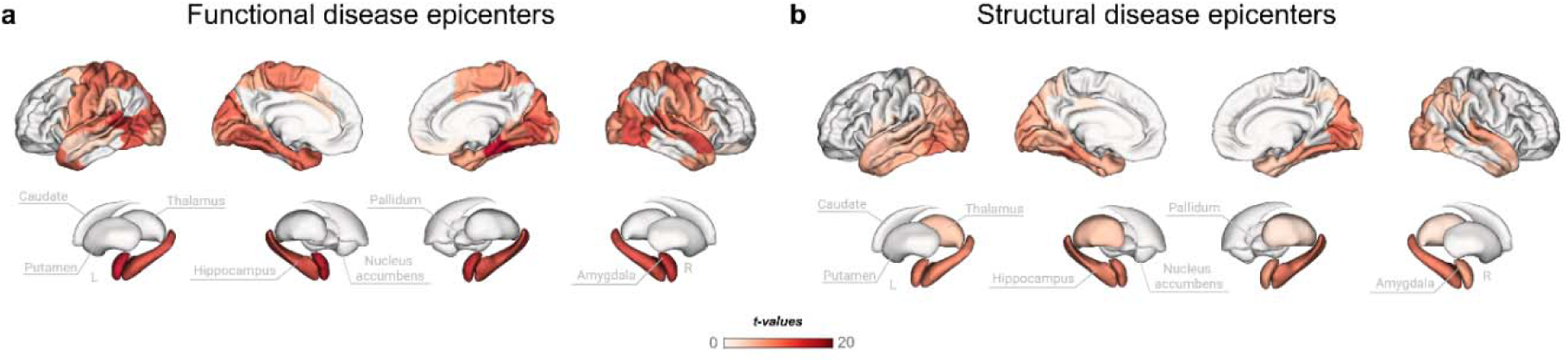
Distribution of structural and functional disease epicenters. Individual-level disease epicenters were computed, Fisher *r*-to-*z* transformed, and assessed for group-level significance using one-tailed *t*-tests with FDR correction. Regions exhibiting significant effects were delineated from non-significant ones. The map displays corresponding *t*-values.

### Findings were robust across a higher-resolution parcellation

In sensitivity analyses using a more fine-grained subdivision of the cortex based on the 400-parcel version of the Schaefer atlas, we showed that our findings were substantially robust across different parcellation resolutions (Figs. S4, and S5).

### A network-based model predicts longitudinal atrophy in individual patients

Finally, we evaluated whether the investigated mechanisms could predict future progression of regional brain atrophy. We modelled longitudinal atrophy progression in each brain region at the group level using linear mixed-effects models with timepoints nested within pwMS/CIS. At the patient level, region-wise estimates of longitudinal atrophy progression were expressed in terms of annualised percentage change (sign-flipped so that more positive values indicated greater atrophy progression) obtained from individual linear models. Patients showed widespread volume/thickness loss over time, more prominent in the temporal, cingulate, and sensorimotor cortices and in subcortical structures (Fig. S6).

We specified generalised additive models (GAMs) using baseline information to predict subsequent atrophy progression in individual patients. We combined data from all pwMS/CIS and all regions to fit a single unified model including non-linear terms for network-based measures (i.e., normative structural strength centrality, normative functional strength centrality, baseline *lesional disconnection* and functional, structural, and transcriptomic neighbourhood atrophy), as well as baseline atrophy, age, follow-up duration, spatial autocorrelation (based on each region’s centre of gravity coordinates), random intercepts for patients and region and a random slope for each region’s relationship between baseline atrophy and subsequent atrophy. The linear fixed effects included sex, site, and a binary variable indicating whether each region was cortical or subcortical. For comparison, we also built a baseline model only including non-network-based predictors.

The network-based model explained 30.3% deviance (adjusted *R^2^*= 0.30) in longitudinal regional atrophy progression, with significant effects of almost all network-based measures. A complete list of predictors, along with their significance and the corresponding interpretation is provided in Table 2, while a graphical depiction of the model terms is available in Fig. S7. Compared to the baseline model not including network measures, the addition of baseline network-based measures significantly enhanced the explanatory power towards subsequent atrophy progression (Akaike information criterion - AIC: 518,785.0 vs 519,108.9, *p* < 001 at the likelihood ratio test).

**Table 2.**
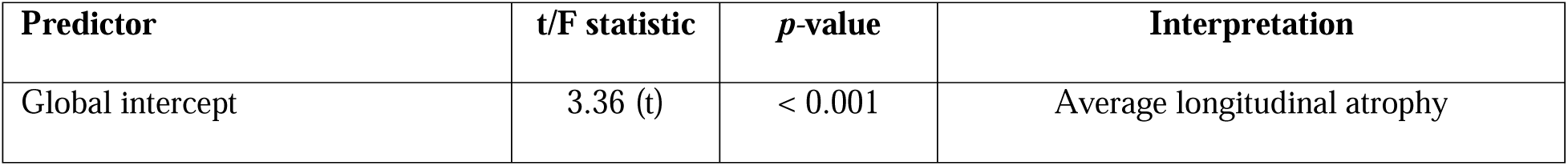
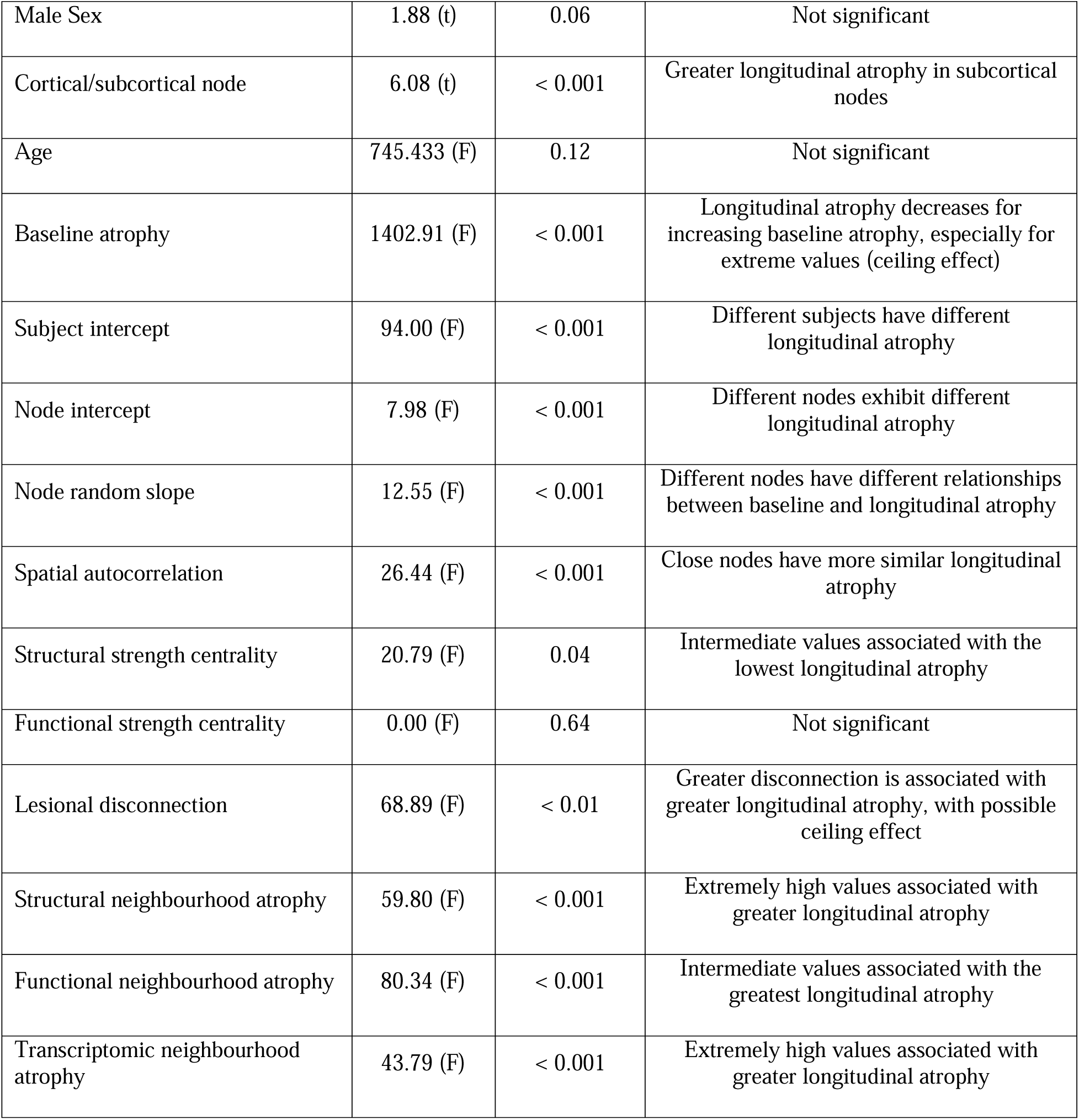
GAM for the prediction of longitudinal atrophy progression. Significance and interpretation of the effect of each predictor on the annualised percentage rate of subsequent longitudinal atrophy.

## Discussion

Using structural MRI from a large multi-centric cohort and a wide set of brain network analyses, we systematically investigated specific hypothesis-driven network-based mechanisms driving GM atrophy in pwMS/CIS. We showed that functional cortical hubs were more susceptible to MS-related atrophy, supporting the *nodal stress* hypothesis. Additionally, *transneuronal degeneration* emerged as a relevant mechanism of neurodegeneration, suggesting that MS-related atrophy might propagate along anatomically and functionally connected regions. A less important role was observed for *lesional disconnection* and *transcriptomic vulnerability*. These network-based mechanisms appeared to be specifically linked to MS-related neurodegeneration rather than to the brain’s physiological structural organisation, and may operate differently in distinct subgroups of pwMS/CIS or phases of the disease. Neurodegeneration patterns are anchored to the connectivity profiles of distinct regions, pointing to the visual, sensorimotor, and temporal cortices, as well as to subcortical structures, as putative disease “epicentres”. Finally, we showed that network-based measures, which are easily obtainable from conventional MRI and normative datasets, could be used to predict future atrophy progression in individual patients.

Functional cortical hubs were associated with greater atrophy, supporting the *nodal stress* hypothesis (*19, 32*). These findings are consistent with prior research showing functional hubness is closely associated with disease vulnerability across conditions, including MS (*16, 33–35*). Regions with high functional centrality, subject to heavy network traffic, may undergo activity-related “wear and tear” that gives rise to or worsens disease-related neurodegeneration, possibly due to high metabolic demands (*33*). In contrast, structural hubs did not show associations with atrophy, which might suggest that highly anatomically connected cortical regions, essential for the integration of brain functioning, are more protected against degeneration through compensatory mechanisms. We also observed that the regional cortical atrophy level was higher for strongly anatomically and functionally connected regions. This finding supports the *transneuronal degeneration* hypothesis, suggesting that neurodegeneration may propagate along brain connections via anterograde and/or retrograde trans-synaptic damage. Such network-based propagation patterns are increasingly recognised in primary neurodegenerative disorders, where pathological changes are thought to spread from regions of greatest/earliest vulnerability (i.e., disease epicentres) to connected areas (*18, 36, 37*). In this context, network architecture has been hypothesised to provide “passive” anatomical conduits through which pathological agents (e.g., misfolded proteins) can be transported (the so-called prion-like hypothesis), as well as to support a more “active” link between altered dynamics of neuronal circuits and neurodegeneration (*36, 38*). Similar mechanisms might also play a role in MS, where the propagation of neuroinflammatory response along WM tracts has also been hypothesised (*39*).

For *lesional disconnection* and *transcriptomic vulnerability*, no associations with regional GM atrophy were observed. Previous studies have investigated the relationship between lesions and atrophy in MS, showing that disruption of WM integrity can lead to trans-synaptic degeneration and, therefore, cortical thinning and subcortical GM volume loss in connected regions (*40*). Similar patterns of secondary GM degeneration following WM damage have also been observed in other neurological disorders, such as stroke and traumatic brain injury disease (*41, 42*). One potential explanation for this apparent discrepancy is that the impact of lesion-induced disconnection on connected GM regions may peak relatively early after the occurrence of a new lesion, to eventually reach a plateau and be superseded by other neurodegeneration mechanisms (*40*). According to this interpretation, the snapshot provided by cross-sectional analyses might not be sensitive enough to detect such an effect, which would be fully exposed only with high-frequency serial acquisition capturing lesion dynamics and subsequent changes in connected GM regions. Also, it should be highlighted that binary lesion masks do not capture lesion heterogeneity and that macroscopic lesions only represent the tip of the iceberg of WM damage in MS. More advanced techniques (e.g., diffusion or quantitative MRI) assessing brain tissue microstructure potentially could provide a more precise account of WM integrity and, consequently, of its impact on regional GM atrophy (*43, 44*). As for the lack of association between normative gene expression profiles and regional GM atrophy, this falls in line with the knowledge of MS as primarily linked to a dysfunction of the immune system rather than to the selective vulnerability of a specific cell population in the central nervous system. However, it should be noted that the normative data is based on a limited number of cases and does not include possible MS-specific alterations in transcriptomic profiles. Hence, future studies focusing on specific genes and/or relying on spatial transcriptomics on MS brain tissues (*25*) will be needed to unravel the spatio-temporal and cell-type-specific gene expression profiles associated with neurodegeneration.

Although the associations between network-based maps and regional atrophy did not reach statistical significance for subcortical regions, they showed similar trends to those observed for cortical areas. This may reflect the fact that subcortical structures exhibit substantial atrophy, potentially diminishing the detectability of network-driven effects, possibly reflecting a floor effect. In addition, the smaller number of subcortical regions compared to cortical regions reduces the statistical power, further limiting the likelihood of significant associations. Moreover, segmentation of subcortical GM using FreeSurfer is known to be less reliable than for cortical GM (*45*), especially in pwMS (*46*), and alternative tools may provide more accurate measures in the future.

Overall, these findings suggest that network-based mechanisms shape MS-related neurodegeneration patterns. Traditionally, MS pathology has been primarily attributed to focal lesions and (acute) inflammation. While current therapies effectively target (acute) inflammatory activity, it is now clear that disease progression can occur independently of relapses and acute inflammation. We speculate that *nodal stress* and *transneuronal degeneration* might play an important role in driving neurodegeneration regardless of lesions, potentially representing treatment targets to tackle progression independent of new lesion activity (*47, 48*).

We then tailored our analyses to individual patients, aiming to assess whether distinct mechanisms affect some pwMS more than others. While the average trends tended to recapitulate the findings of the group-level analyses, with regional GM atrophy most strongly associated with that of connected neighbours, individual-level analyses revealed a specific degree of inter-subject variability which was not discernible at the group level, especially for subcortical regions. This may reflect greater biological heterogeneity in the mechanisms driving subcortical neurodegeneration in MS, although methodological reasons such as the greater measurement error associated with smaller brain structures (*45, 49*) plus the lower stability of correlations drawn from a smaller number of regions, might also play a role.

When looking at data-driven atrophy subgroups, we identified two distinct patterns: one characterised by widespread subcortical and cortical neurodegeneration, and the other showing predominantly subcortical atrophy with relative sparing of the cortex. Patients assigned to the two subgroups exhibited similar disease durations, as well as comparable proportions of clinical phenotypes, suggesting that the observed GM atrophy profiles don’t represent consecutive phases of the disease but rather two distinct “modes” of neurodegeneration, which substantially align with previously described MRI-driven subtypes (*5, 50*). When comparing hypothesised neurodegeneration drivers across MRI-driven and clinical phenotypes, significant differences emerged, with network-based mechanisms apparently more relevant in patients with widespread atrophy and more advanced disease phases. Taken together, results of the subgroup-level analyses seem to suggest that the hypothesised network-based mechanisms are specifically associated with MS-related neurodegeneration, rather than reflect the physiological organisation of brain structure (i.e., their associations with regional volume/thickness only become apparent in the presence of disease-related atrophy), and may have varying impact according to the phase and phenotype of the disease.

We also tested the hypothesis that the connectivity profiles of specific regions constrain the observed atrophy patterns. We identified candidate structural and functional epicentres in both cortical and subcortical regions, from which the propagation of neurodegeneration to connected regions supposedly starts. Cortical epicentres were primarily located in the visual and temporal areas, as well as in the sensorimotor cortex, possibly reflecting regions of high connectivity that may facilitate network-level spread of atrophy. Subcortical epicentres included the hippocampus, amygdala, and thalamus. These findings can be interpreted in light of existing knowledge on MS. For instance, involvement of optic pathways, including secondary atrophy of the visual cortex, is a common early manifestation of the disease (*51*), as well as thalamic atrophy (*52*). It is important to note that our epicentre mapping analysis mostly assumes *transneuronal degeneration* as the mechanism driving neurodegeneration, which might not be fully accurate. While the observed epicentres provide a meaningful approximation, more complex models taking into account other mechanisms and/or incorporating longitudinal assessments are warranted to fully understand how neurodegeneration propagates across the brain.

Longitudinal modelling showed that network-derived metrics significantly enhance the prediction of atrophy progression beyond baseline damage and demographic factors. Baseline atrophy was a significant predictor of subsequent atrophy progression, which decreased for increasing baseline atrophy, especially for extreme values, possibly reflecting a ceiling effect of structural brain damage. Along with spatial autocorrelation (i.e., close regions have more similar longitudinal atrophy) and subcortical vulnerability (i.e., subcortical regions exhibit greater longitudinal atrophy), all network-based regional properties, except for functional network centrality, emerged as significant predictors of longitudinal atrophy. Notably, *lesional disconnection* and neighbourhood atrophy measures, which are (partially) derived from patient-level data, were more significantly associated with longitudinal atrophy progression than network centrality measures, which are purely based on normative connectomes. This suggests that patient-specific network information might be more strongly linked to future atrophy progression, warranting future studies using advanced MRI to obtain individual-level structural and functional connectomes (*53*).

Several additional limitations should be acknowledged. First, although our cohort included data from multiple European centres, disease duration was relatively long on average, meaning that the earliest stages of MS were underrepresented. Second, we analysed cortical and subcortical regions separately in order to mitigate the confounding effects of spatial autocorrelation, but this choice limited our ability to fully characterise the entire brain network and subcortico-subcortical interactions, which may also play an important role in disease propagation. Finally, functional and diffusion MRI data were not available for all pwMS, as well as transcriptomic data, which led us to use normative connectomes and gene expression profiles. Future studies should aim to incorporate actual data from individual pwMS to better reflect disease-specific characteristics and their heterogeneity across participants.

MS-related GM atrophy is shaped by network architecture, with *nodal stress* and *transneuronal degeneration* emerging as central mechanisms of neurodegeneration among the mechanisms investigated. Visual, sensorimotor and temporal cortices, as well as subcortical structures including the hippocampi and thalami, may act as epicentres of neurodegeneration. Network-based measures, which are easily obtainable from conventional MRI and normative datasets, can help predict future atrophy progression in individual patients. These findings provide a mechanistic framework for understanding MS-related neurodegeneration, potentially informing patient prognostication and therapeutic approaches.

## Materials and Methods

Unless otherwise specified, statistical analyses were performed in R (v4.4.1, R Foundation).

### Participants

In this retrospective multicentric study, we collected structural brain MRI and clinico-demographic data of patients diagnosed with MS according to the 2017 McDonald criteria (*54*) or CIS (*55*), as well as of HCs. Included participants were part of the Amsterdam (NL74887.029.20), Prague, and Graz MS cohorts (*56, 57*). Exclusion criteria were age younger than 18 or older than 75 years, the presence of other relevant neurologic, psychiatric, or systemic conditions, and having had relapses or received steroid treatment within the month preceding the visit. Written informed consent was obtained from each participant independently at each centre following standard procedures.

### MRI acquisition and processing

Participants underwent longitudinal brain MRI on 3T scanners including 3D T1w and 3D FLAIR sequences (Table S1). T2-hyperintense lesions were automatically segmented on FLAIR and T1w scans using LST-AI (*58*), and the corresponding masks were used to fill lesions in T1w images through NiftySeg’s lesion filling procedure (*59*).

Lesion-filled T1w volumes were processed with FreeSurfer (v7.4.1) using the recon-all cross-sectional and longitudinal pipelines to obtain thicknesses of 100 cortical regions from the Schaefer atlas (*26*) and volumes of 14 subcortical structures from FreeSurfer’s Aseg atlas (*27*). To ensure high quality of the included scans, we used the Euler number, an index of cortical surface reconstruction quality generated automatically by FreeSurfer. In particular, we discarded scans below an Euler number cutoff threshold of −120 (*60*).

In addition, disconnection matrices were estimated from lesion masks and the HCP1065 tractography atlas using the Lesion Quantification Toolkit (LQT) (*28*). Briefly, individual lesion masks were registered to the MNI space by applying the nonlinear transformation obtained by normalizing T1w volumes to the template using ANTs v2.5.1. Then, based on spatially normalised lesion masks, the pairwise disconnection between pairs of GM regions was computed as the proportion of streamlines intersecting lesions and used to fill subject-level 114 × 114 structural disconnection matrices.

To assess the stability of the findings across different parcellation resolutions, we also conducted sensitivity analyses using a more fine-grained subdivision of the cortex based on the 400-parcel version of the Schaefer atlas (*26*).

### Atrophy mapping

To quantify regional atrophy at baseline, we generated W-score maps for each participant, following an established procedure (*18, 61*). Region-wise linear regressions were first performed in the HC group to estimate the physiological effects of age, sex, intracranial volume (for subcortical volumes), and site on cortical thickness and subcortical volume values. W-score maps were then computed for each participant using the following formula:

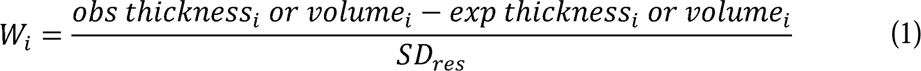

where *W_i_* is the W-score at region *i*, *obs thickness*_i_ or v*olume*_i_ is the observed cortical thickness or subcortical volume at region *i*, *exp thickness*_i_ or v*olume*_i_ is the expected cortical thickness in matched HCs at region *i* given the subject’s age, sex, intracranial volume (for subcortical volumes), and site provenance (*thickness/volume* ∼ *age* + *sex* [+ *intracranial volume*] + *site*), and SD*_res_* is the standard deviation of the residuals in the HCs group. For interpretability, W-scores were inverted in sign such that more positive scores indicated greater atrophy whereas more negative scores denoted lower atrophy. Group-level cross-sectional atrophy was expressed as the effect size (Cohen’s *d*) of the difference between W-score maps of pwMS/CIS and HCs, assessing statistical significance with a *t*-test and adjusting for multiple comparisons using false discovery rate (FDR)-correction, with *p* < 0.05 indicating statistical significance.

### Spatial permutation tests

We used the ENIGMA Toolbox (v2.0.3, https://github.com/MICA-MNI/ENIGMA) in Python v3.9.0 to perform spatial correlations. As the intrinsic spatial smoothness in two given brain maps is known to potentially inflate the significance of their spatial correlation, we assessed statistical significance of spatial correlations against spatial autocorrelation-preserving null models (*62*). In particular, for cortical regions, we used spin permutation tests, where null models of overlap between cortical maps are generated by projecting the spatial coordinates of cortical data onto the surface spheres, applying randomly sampled rotations (10,000 repetitions), and reassigning regional values (*62*). The observed correlation coefficients are then compared against the null distributions determined by the ensemble of correlation coefficients comparing spatially permuted cortical maps. To compare spatial overlap between subcortical maps, we used a similar approach with the exception that subcortical labels were randomly shuffled as opposed to being projected onto spheres (*16*).

### Network-based atrophy modelling

To investigate the different mechanisms hypothesised to underlie MS-related neurodegeneration, we obtained a set of network-based maps using normative or participants’ data, and assessed their spatial similarity with group-level regional atrophy. For connectivity-based mechanisms, we relied on normative structural (SC) and functional (FC) connectomes derived from 206 healthy adults sampled from the Human Connectome Project (HCP) (*54, 63*) and made available by the ENIGMA toolbox (*64*). For transcriptomic analyses, we used gene expression data provided by the Allen Human Brain Atlas (AHBA), a brain-wide gene expression atlas obtained from six human brain donors comprising microarray-derived measures from over 20,000 genes. Gene expression data, processed according to established recommendations (*15*) including mapping onto the Schaefer parcellations and removal of genes whose similarity across donors fell below a *r* < 0.2 threshold (leaving a total of 11,053 genes for the 100-parcel and 7,584 for the 400-parcel versions), were obtained through the ENIGMA toolbox. Finally, we correlated the rows of the region × gene expression matrix in pairs to generate a symmetric region × region correlated gene expression (CGE) matrix, indicating the similarity of gene expression profiles between different brain regions (*15*).

To investigate the *nodal stress* hypothesis, we calculated regional strength centrality maps from the normative structural and functional connectomes as the sum of all cortico-cortical or subcortico-cortical edges values insisting on each node, and tested their spatial correlation with GM atrophy (*16*).

For *transneuronal degeneration*, we tested the spatial correlation between cortical and subcortical atrophy and neighbourhood atrophy maps, calculated as follows:

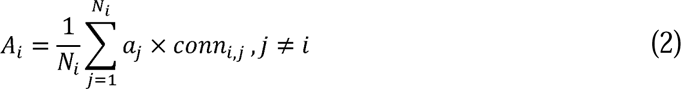

where *A_i_* is the average neighbourhood atrophy of node *i*, *a_j_* is the atrophy of the *j*-th neighbour of node *i*, *conn_i,j_* is the strength of connection between nodes *i* and *j*, and *N_i_* is the total number of node *i*’s first-degree neighbours. Self-connections (i.e., of a node with itself) were excluded (*j*≠*i*), while connectivity *conn_i,j_*was defined according to normative structural or functional connectomes in separate analyses (*18*). As a result, we obtained functional and structural neighbourhood atrophy maps.

To investigate whether atrophy was associated with lesion-induced disconnection, we computed a node disconnection map as the regional strength centrality (i.e., the sum of all cortico-cortical or subcortico-cortical edges insisting on each node) of the data-driven disconnectome matrix, and tested its spatial correlation with GM atrophy.

Finally, for *transcriptomic vulnerability*, we tested the spatial correlation between GM atrophy and transcriptomic neighbourhood atrophy maps, calculated as in (2) but using transcriptomic similarity defined according to the normative CGE matrix (instead of connectivity) as the weighting factor.

### Patient- and subgroup-tailored atrophy modelling

Network-based mechanisms were also assessed in individual patients and specific subgroups. In particular, for each patient, we assessed the spatial similarities between the individual W-score map and network-based maps, thus obtaining a correlation coefficient (*r*) and a *p*-value (from spin/shuffle permutation tests with 10,000 repetitions) expressing the relevance of each hypothesised mechanism on a per-subject basis. To enable comparisons across subjects and subgroups, we transformed *r* values into Fisher’s *z* values using the inverse hyperbolic tangent function (*z* = arctanh(*r*)).

After assessing the distributions of each mechanism’s relevance across subjects, we investigated whether they differentially contributed to the observed atrophy patterns across distinct MRI-driven and clinically-defined patients’ subgroups. Specifically, we subgrouped participants into distinct regional atrophy (i.e., W-score map) profiles using the HYDRA algorithm, a non-linear semi-supervised machine learning approach for simultaneous binary classification (patients vs controls) and subtype identification (*29*). We tested different numbers of clusters (from *k*=1 to *k*=5) with default parameters, and assessed the clustering performance by taking into account the stability of the obtained results: we used the Adjusted Rand Index (ARI) to quantify the similarity between clustering results across training folds in a 100-times repeated 10-fold cross-validation scheme (*29*). The clustering corresponding to the highest ARI was selected for subsequent analyses. Alternatively, we subgrouped patients based on their clinical phenotypes as early relapsing (ER, CIS and RRMS with < 5 years disease duration (DD)), late relapsing (LR, RRMS with ≥ 5 years DD), SPMS, and PPMS (*30*). Differences in terms of *z*-values, from zero and between subgroups, were assessed in a linear model framework, correcting for multiple comparisons across mechanisms and subgroup pairs using FDR.

### Mapping disease epicentres

To identify potential disease epicentres of neurodegeneration, we performed patient-tailored connectivity-based epicentre mapping following a previously described approach (*31*). We hypothesised that an epicentre would be a region whose intrinsic connectivity pattern most strongly resembled the patient’s atrophy profile. In this light, we systematically compared seed-based (across all regions) normative structural and functional connectomes with individual atrophy (i.e., W-score) maps. As a result, we obtained structural and functional epicentre scores (*r*-values) on a per-patient and per-region basis. After Fisher’s *r*-to-*z* transformation, we determined the significance of candidate epicentres at the group level with a one-sample, one-tailed *t-*test, correcting for multiple comparisons across brain regions using FDR. The resulting statistical *t* maps (and corresponding *p* values) represented inferred structural and functional epicentre degrees across parcels at the group level.

### Network-based prediction of atrophy progression

Longitudinal atrophy progression was modelled in patients with at least two MRI visits. At the group level, linear mixed effects models were used to examine the change in cortical thickness or subcortical volume across follow-up visits: *thickness* or *volume* ∼ 1 + *time* + (1 + *time* | *subject*). Time since baseline visit (years) was used as fixed effect of interest, while subject-wise slope and intercept were modelled as random effects. Separate models were fitted for each parcel, with the resulting *t* map corresponding to the region-wise effect of time on cortical thickness/subcortical volume. Subject-level regional atrophy rates were estimated with individual linear models and expressed as the slope of regional thickness/volume change over time, then converted to annualised percentage change. In the same way as for cross-sectional atrophy, longitudinal estimates were multiplied by −1 such that more positive scores indicated greater atrophy progression whereas more negative scores denoted lower atrophy progression.

Using baseline information, we specified a model to predict subsequent atrophy progression in individual patients. We used generalised additive models (GAMs), as implemented in the *mgcv* package’s *bam* function, combining data from all patients and all regions to fit a single unified model. GAMs are well suited for modelling spatiotemporal change because they can fit predictors in a flexible, non-linear fashion with safeguards against overfitting (*65*). These models can also include linear terms, random effect terms, and adjustment for spatial correlation among neighbouring regions. Our model included non-linear terms for network-based measures (i.e., normative structural strength centrality, normative functional strength centrality, baseline lesional disconnection and functional, structural, and transcriptomic neighbourhood atrophy), as well as baseline atrophy, age, follow-up duration, spatial autocorrelation (based on each region’s centre of gravity coordinates), random intercepts for patients and region and a random slope for each region’s relationship between baseline atrophy and subsequent atrophy. The linear fixed effects were sex, site, and a binary variable identifying each region as cortical or subcortical. We used the fast REML estimation method, enabling selection penalties to be added to the smooth effects. To assess the added value of network-derived metrics in predicting subsequent atrophy, we also fitted a baseline model including only non network-based predictors. Comparison was made in terms of the models’ Akaike Information Criterion (AIC) and with a Likelihood Ratio Test (LRT).

## Acknowledgments

We gratefully acknowledge all the individuals who participated in this research, including people with multiple sclerosis, caregivers, and volunteers, whose contributions made this work possible and helped advance our understanding of multiple sclerosis.

## Funding

This study is sponsored by ZonMW VIDI grant 09150172010056 (MMS)

## Author contributions

Conceptualization: MBC, MT, LL, GP

Data curation: MBC, MT, DRvN, IK, LL, BH, SR, DP, CE, TU, MV, JK, EMMS, MDS, HV, FB, MMS and GP

Methodology & Investigation: MBC, GP

Supervision: MMS, GP

Writing—original draft: MBC, GP

Writing—review & editing: MBC, MT, DRvN, IK, LL, BH, SR, DP, CE, TU, MV, JK, EMMS, MDS, HV, FB, MMS and GP

## Competing interests

MBC is supported by research grants from Merck and Atara Biotherapeutics.

DRvN is supported by the European Union’s Horizon Widera programme under grant agreement no. 101159624 (TACTIX).

IK received research grants from LabEx TRAIL (Translational Research and Advanced Imaging Laboratory) and ARSEP (Fondation pour l’Aide à la Recherche sur la Sclérose En Plaques) and speakers’ honoraria from Celgene.

LL receives funding from the MSCA postdoctoral fellowship (#101204296).

BH has received speaker honoraria from Roche, Bristol-Myers Squibb, and Sanofi, research funding from ECTRIMS, and travel funding from Janssen.

DP is a member of the advisory board for “Cognition and MS” for Novartis and has received speaking honoraria from Biogen, Novartis, MedAhead and Bristol-Myers Squibb.

CE has received funding for traveling and speaker honoraria from Biogen Idec, Bayer Schering Pharma, Merck Serono, Novartis, Genzyme and Teva Pharmaceutical Industries Ltd./sanofi-aventis, Shire; received research support from Merck Serono, Biogen Idec, and Teva Pharmaceutical Industries Ltd./sanofi-aventis; and serves on scientific advisory boards for Bayer Schering Pharma, Biogen Idec, Merck Serono, Novartis, Genzyme, Roche, and Teva Pharmaceutical Industries Ltd./sanofi- Aventis.

TU received financial support for conference travel and honoraria from Biogen, Novartis, Roche, Bristol Myers Squibb, and Merck, as well as support for research activities from Biogen and Sanofi. He also received support by the Czech Ministry of Health, the institutional support of a hospital research project (MH CZ-DRO-VFN64165), the Czech Ministry of Health project (NU22-04-00193) and the Charles University Cooperation Program in Neuroscience.

MV received speaker honoraria and consultant fees from Biogen, Novartis, Roche, Sanofi, and Teva, as well as support for research activities from Biogen. She also received support by the Czech Ministry of Health, the institutional support of a hospital research project (MH CZ-DRO-VFN64165), the Czech Ministry of Health project (NU22-04-00193) and the Charles University Cooperation Program in Neuroscience.

JK received research grants for multicentre investigator-initiated trials DOT-MS trial, ClinicalTrials. gov Identifier: NCT04260711 (ZonMW) and BLOOMS trial (ZonMW and Treatmeds), ClinicalTrials. gov Identifier: NCT05296161); received consulting fees for F. Hoffmann-La Roche, Biogen, Teva, Merck, Novartis and Sanofi/Genzyme (all payments to institution); reports speaker relationships with F. Hoffmann-La Roche, Biogen, Immunic, Teva, Merck, Novartis and Sanofi/Genzyme (all payments to institution); adjudication committee of MS clinical trial of Immunic (payments to institution only).

EMM serves on the editorial board of Neurology and Frontiers in Neurology and received research support from Stichting MS Research and ZonMW. Received speaker fees from Merck and Novartis.

HV has received research support from Merck, Novartis, Pfizer, and Teva, consulting fees from Merck, and speaker honoraria from Novartis; all funds were paid to his institution.

FB serves on the steering committee and is iDMC member for Biogen, Merck, Roche, EISAI, acts as a consultant for Roche, Biogen, Merck, IXICO, Jansen, Combinostics, has research agreements with Novartis, Merck, Biogen, GE, Roche and is co-founder and shareholder of Queen Square Analytics LTD.

MMS serves on the editorial board of Neurology, Multiple Sclerosis Journal and Frontiers in Neurology, receives research support from the Dutch MS Research Foundation, Eurostars-EUREKA, ARSEP, Amsterdam Neuroscience and ZonMW (Vidi grant, project number 09150172010056) and has served as a consultant for or received research support from Atara Biotherapeutics, Biogen, Celgene/Bristol Meyers Squibb, EIP, Sanofi, MedDay and Merck.

GP has received research grants from ECTRIMS, MAGNIMS, ESNR, and ZonMW.

All other authors declare they have no competing interests.

## Data and materials availability

Study data are available from the corresponding author upon reasonable request, subject to privacy and ethical guidelines. The code used for the analyses is available on GitHub (https://github.com/giupontillo/AtrophyModellingMS). All additional resources are cited in the main text or the supplementary materials.

## Supplementary Materials

**Fig. S1.**
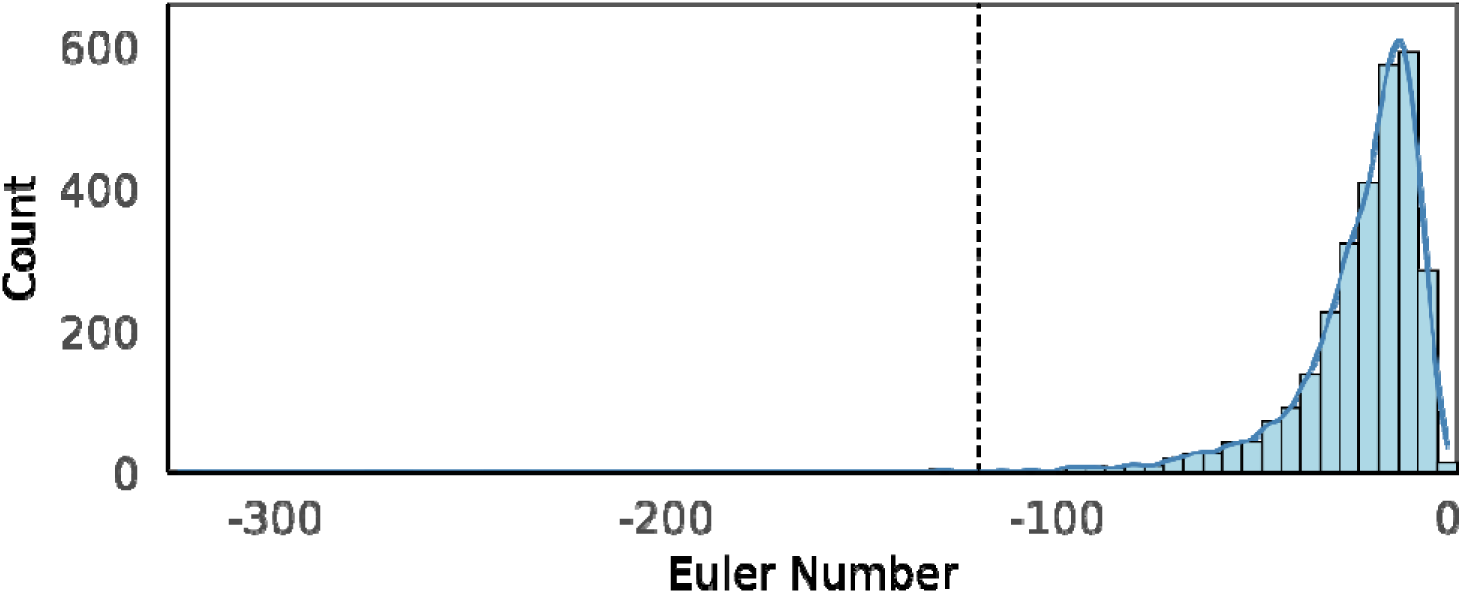
Histogram showing the distribution of Euler numbers for all processed scans, with the dashed line placed at the cutoff threshold of −120.

**Fig. S2.**
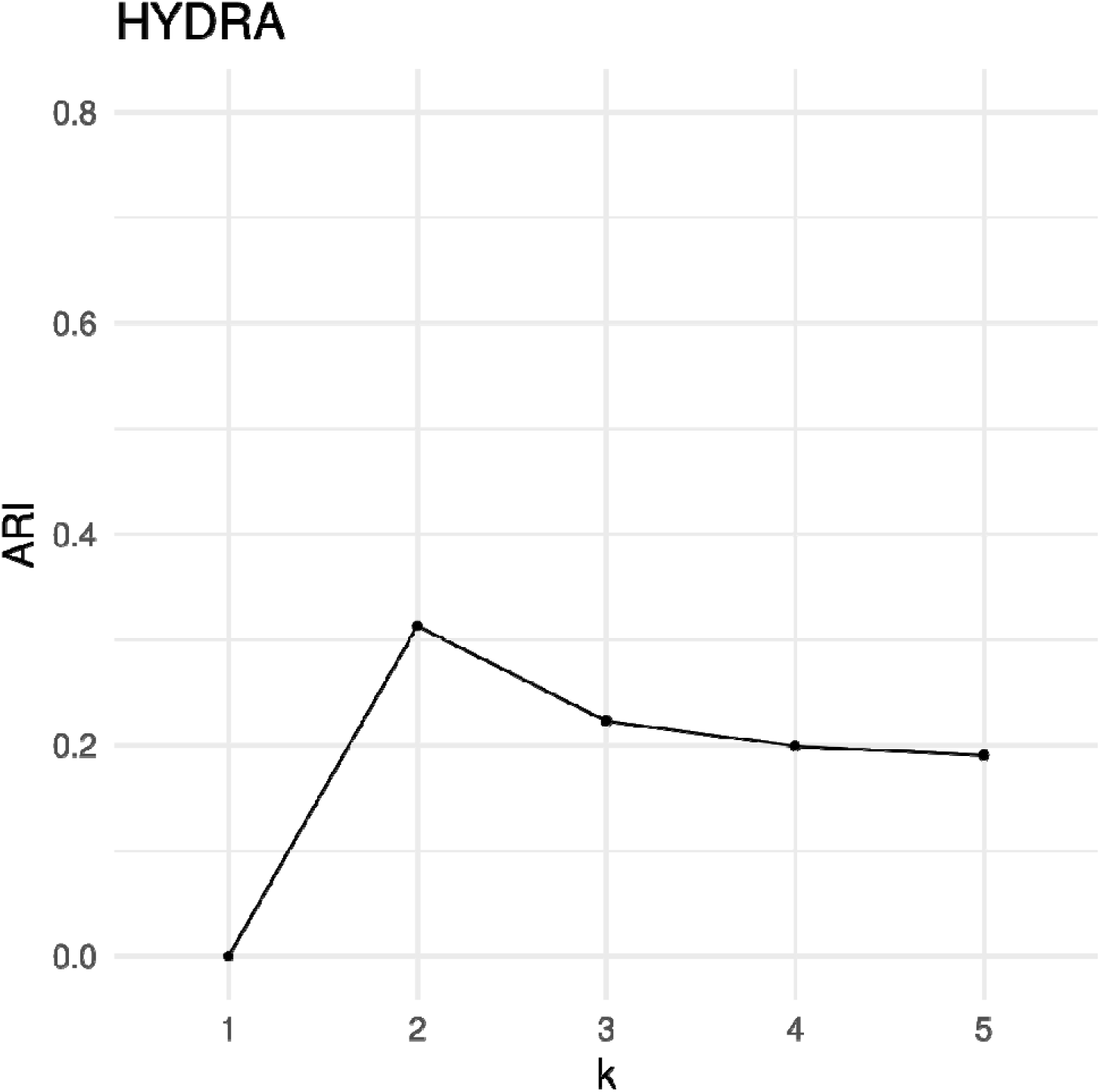
Results of the data-driven clustering using HYDRA. Plot showing the stability of the identified clusters across cross-validation folds, expressed in terms of Adjusted Rand Index (ARI). We tested different numbers of clusters (from *k* = 1 to *k* = 5), with the cluster stability maximized at *k* = 2.

**Fig. S3.**
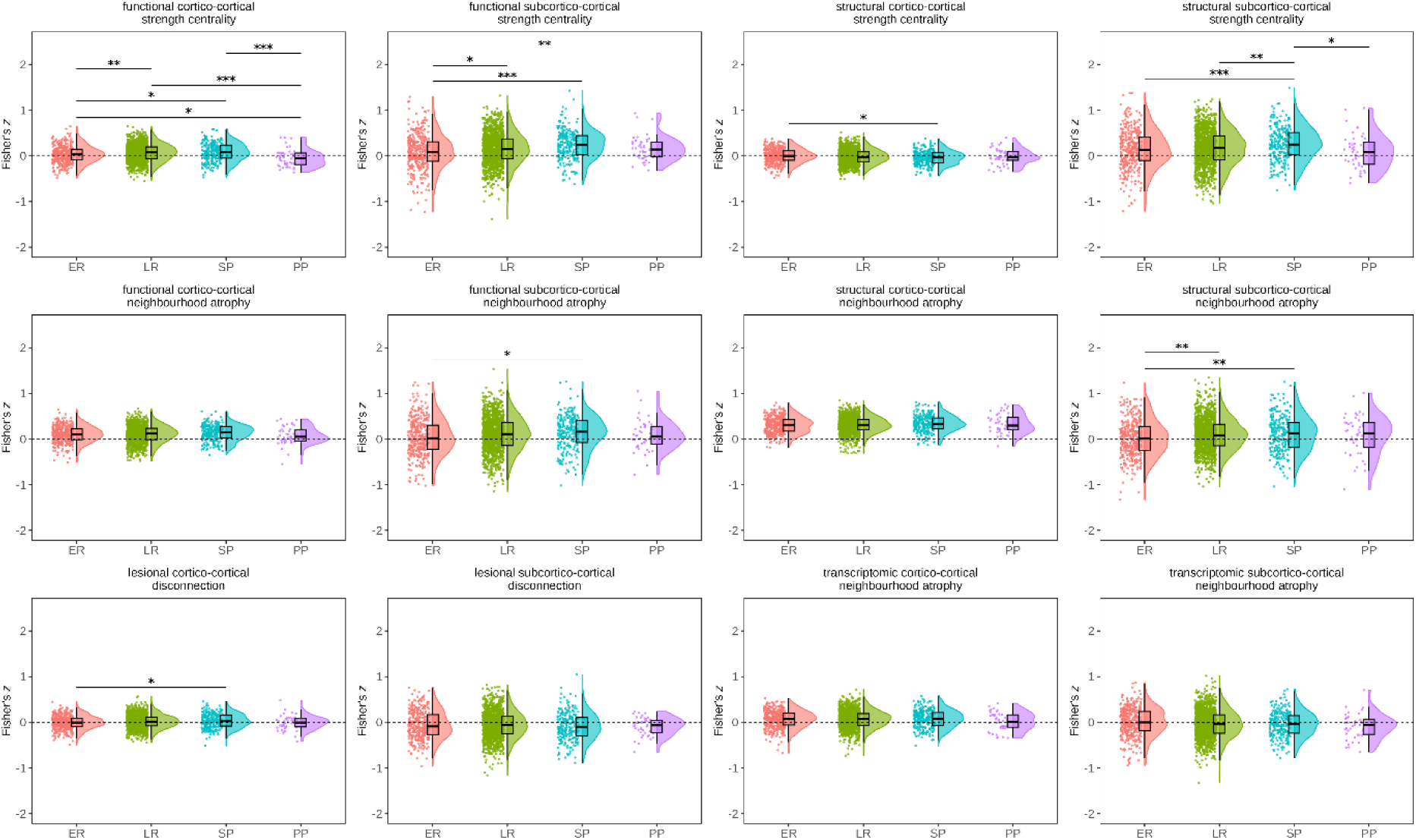
Putative neurodegeneration mechanisms across clinical phenotypes. Differences between clinical phenotypes were first identified using ANOVA, followed by Tukey’s post-hoc pairwise comparisons. *p* < 0.05 (*), *p* < 0.01 (**), *p* < 0.001 (***). Abbreviations: ER = early relapsing; LR = late relapsing; PP = primary progressive; SP = secondary progressive.

**Fig. S4.**
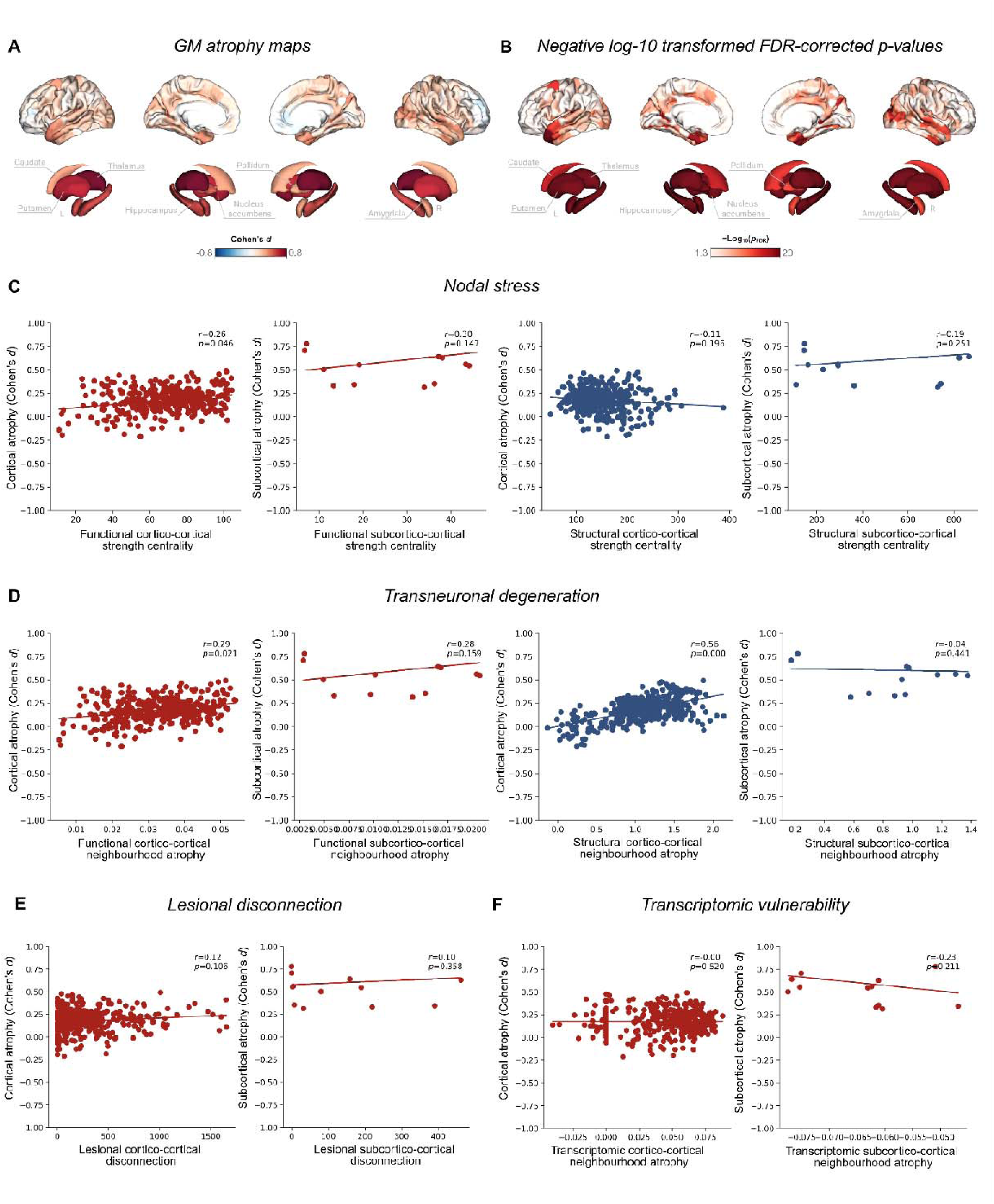
Network-based atrophy mechanisms using Schaefer 400 parcellation. (A) Atrophy maps show cross-sectional differences (Cohen’s d) in cortical thickness and subcortical volumes between pwMS/CIS and HCs, adjusting for physiological effects of age, sex, intracranial volume (for subcortical volumes), and site. (B) Negative log10-transformed FDR-corrected p-values are shown. (C) Correlation between functional and structural strength centrality and grey matter atrophy. (D) Correlation between functional and structural neighbourhood atrophy and grey matter atrophy. (E) Correlation lesion-related disconnection and (F) transcriptomic neighbourhood atrophy and grey matter atrophy.

**Fig. S5.**
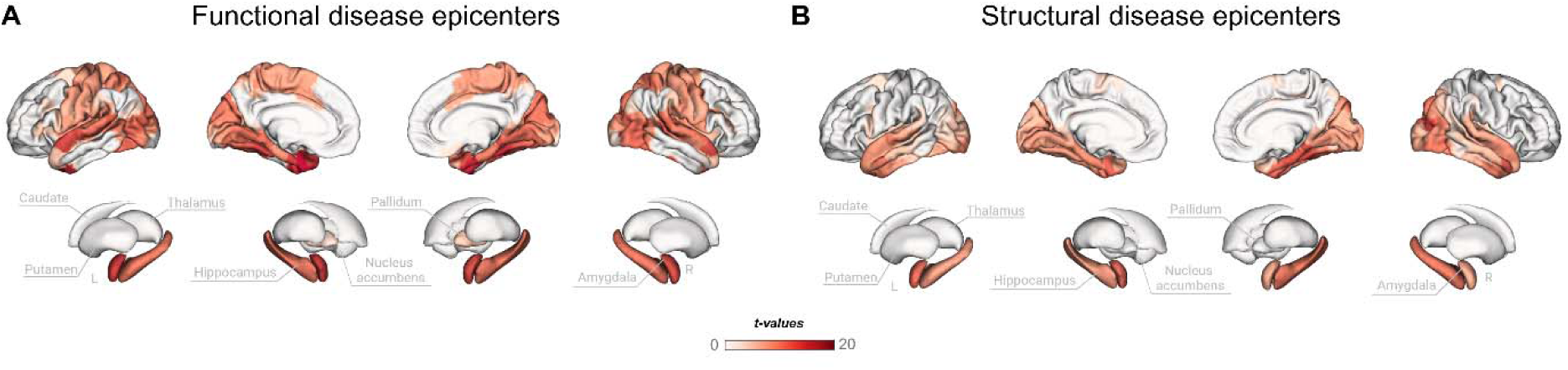
Disease epicentres using Schaefer 400 parcellation. Individual-level disease epicentres were computed, Fisher *r*-to-*z* transformed, and assessed for group-level significance using one-tailed *t*-tests with FDR correction. Shown are regions exhibiting significant effects, with maps displaying the corresponding *t*-values.

**Fig. S6.**
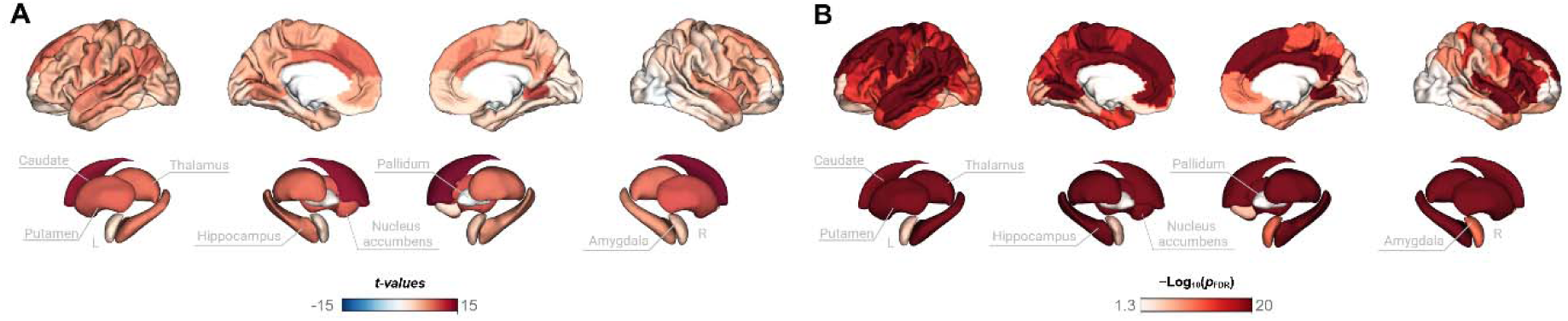
Longitudinal atrophy profiles. (A) Atrophy maps show longitudinal atrophy (*t*-values) in cortical thickness and subcortical volumes. Longitudinal atrophy progression was modelled in patients with at least two MRI visits. (B) Negative log10-transformed FDR-corrected *p*-values are shown.

**Fig. S7.**
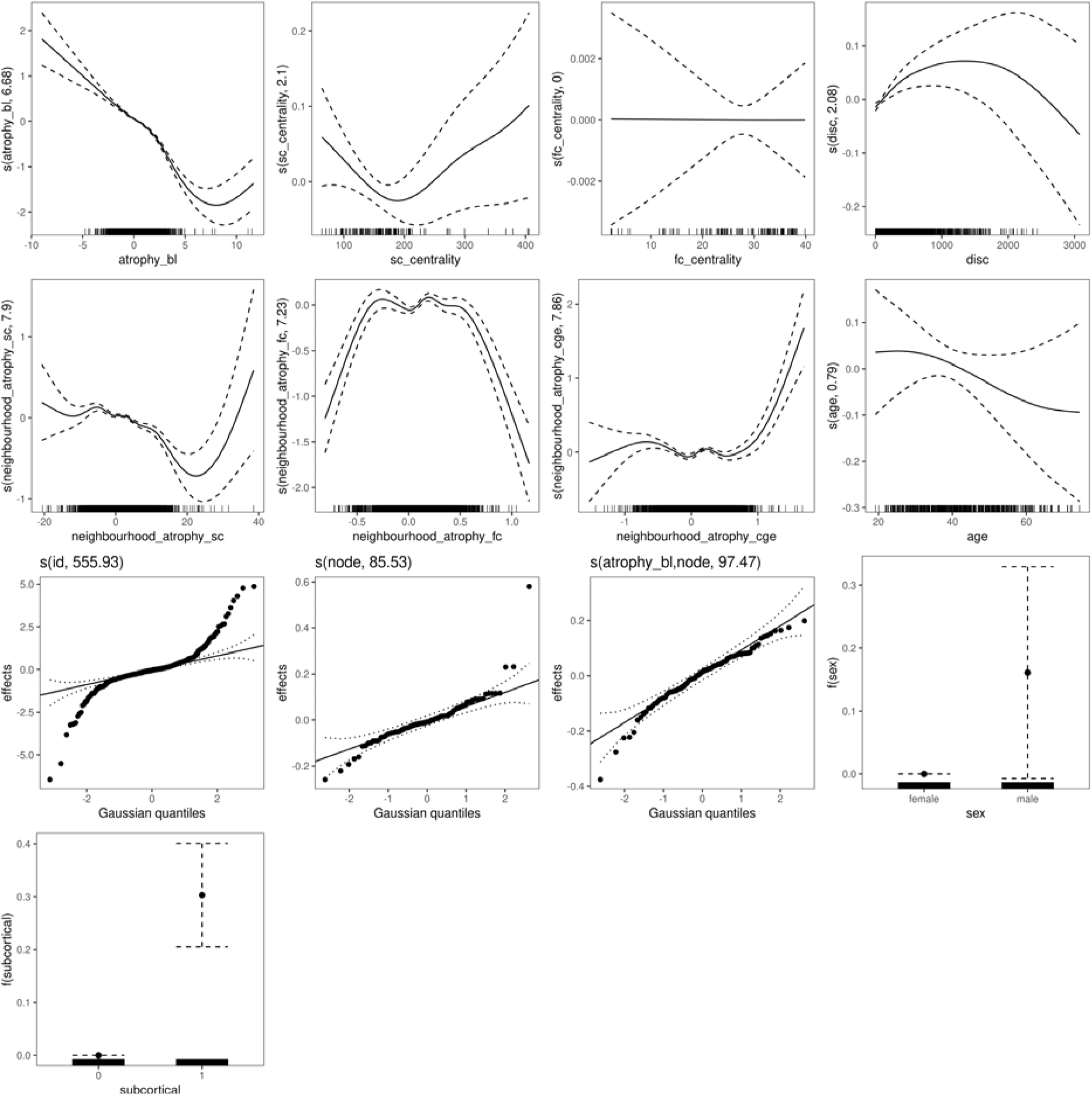
Network-based model for predicting longitudinal atrophy. Effect plots for the predictors of interest, where dashed lines correspond to 95% confidence interval of the fit.

**Table S1.** MRI acquisition protocols. Abbreviation: FLAIR = fluid-attenuated inversion recovery; mm = millimetre; ms = milliseconds; T1w = T1-weigthed; TE = echo time; TI = inversion time; TR = repetition time; FA = flip angle; sag = sagittal.

| Centre | Amsterdam |  | Graz |  | Prague |  |
| --- | --- | --- | --- | --- | --- | --- |
| Field strength | 3 Tesla |  | 3 Tesla |  | 3 Tesla |  |
| Vendor | GE |  | Siemens |  | Siemens |  |
| Model | Discovery MR750 |  | Prisma |  | Skyra |  |
| Years | 2014-2023 |  | 2021-2022 |  | 2012-2022 |  |
| Modality | **T1w** | **FLAIR** | **T1w** | **FLAIR** | **T1w** | **FLAIR** |
| Voxel dimensions (mm<sup>3</sup>) | 0.94x0.94x1 | 0.98x0.98x1.2 | 1x1x1 | 1x1x1 | 1x1x1 | 1x1x1 |
| TR (ms) | 7.8 | 8000 | 1900 | 5000 | 2300 | 5000 |
| TE (ms) | 3 | 125 | 2.7 | 393 | 3 | 397 |
| TI (ms) | 450 | 2350 | 900 | 1800 | 900 | 1800 |
| FA (°) | 12 | - | 9 | 120 | 9 | 120 |
| Slices, Orientation | 172, sag | 128, sag | 176, sag | 176, sag | 176, sag | 176, sag |

**Table S2.**
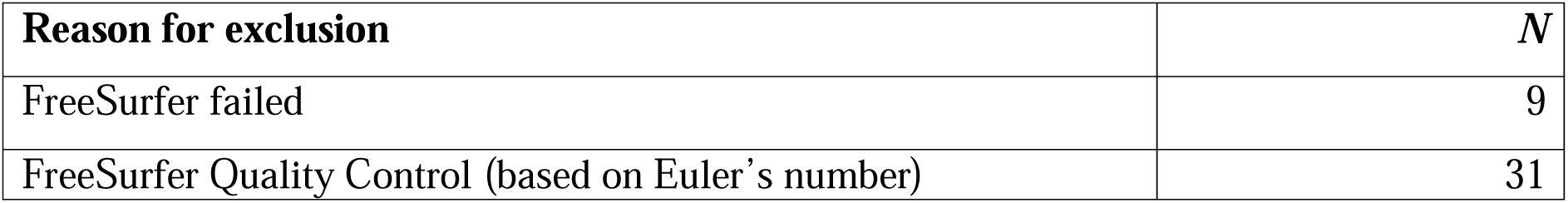
Number and reason for excluded scans. Exclusion reasons are summarized for all participants who were not included in the final analysis. Numbers represent the count of sessions excluded for each reason.

**Table S3.**
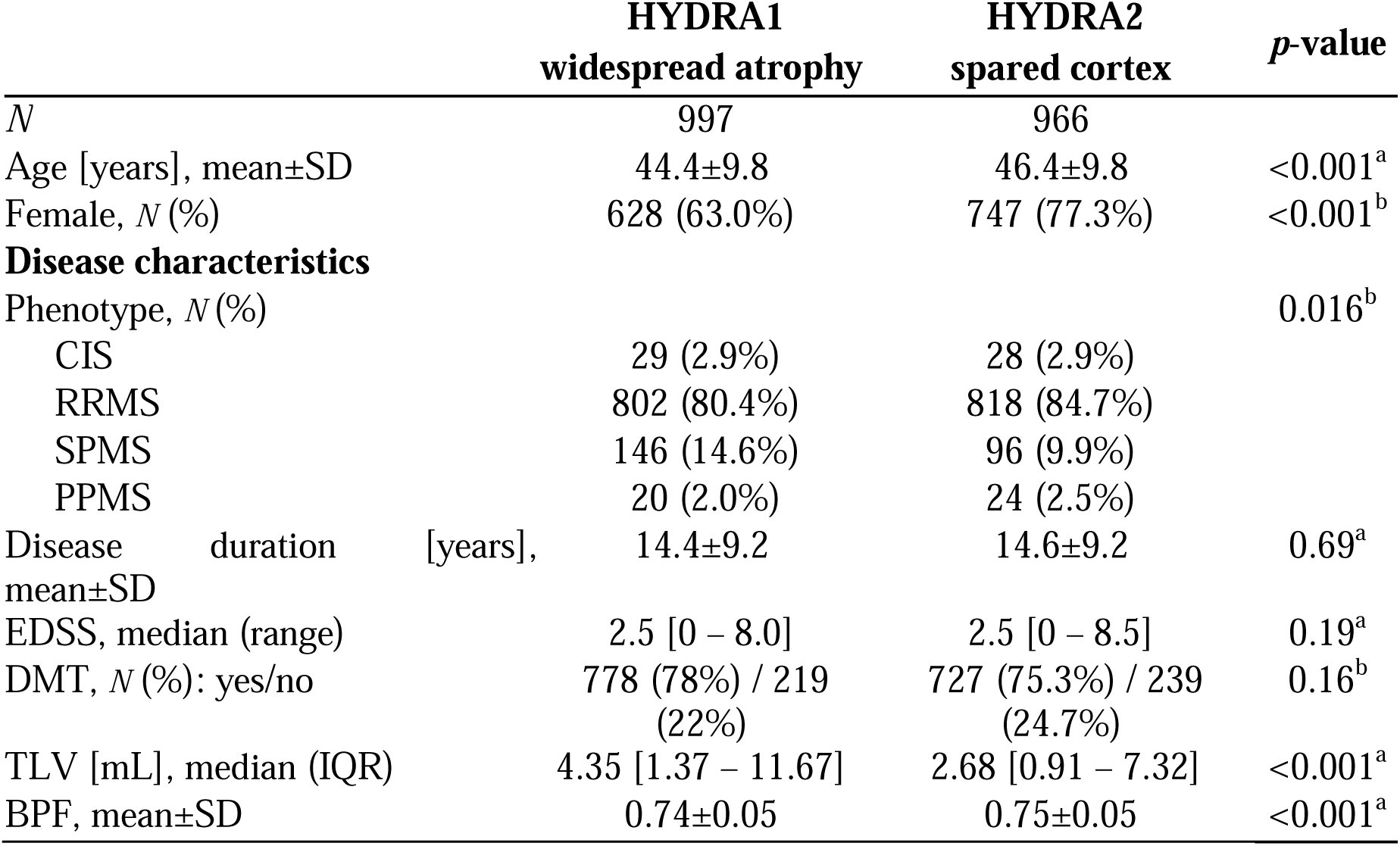
Demographic, clinical and MRI characteristics of HYDRA subgroups. Baseline characteristics of included data-driven HYDRA groups. Abbreviations: BPF = brain parenchymal fraction, CIS = clinically isolated syndrome, DMT = disease-modifying treatment, EDSS = Expanded Disability Status Scale, IQR = interquartile range, mL = millilitres, PPMS = primary-progressive multiple sclerosis, RRMS = relapsing-remitting multiple sclerosis, SD = standard deviation, SPMS = secondary-progressive multiple sclerosis, TLV = total lesion volume. ^a^Permutation *t*-test (n = 10,000); ^b^Chi-square test.

## Notes

### Competing Interest Statement

MBC is supported by research grants from Merck and Atara Biotherapeutics.
DRvN is supported by the European Unions Horizon Widera programme under grant agreement no. 101159624 (TACTIX).
IK received research grants from LabEx TRAIL (Translational Research and Advanced Imaging Laboratory) and ARSEP (Fondation pour lAide a la Recherche sur la Sclerose En Plaques) and speakers honoraria from Celgene.
LL receives funding from the MSCA postdoctoral fellowship (#101204296).
BH has received speaker honoraria from Roche, Bristol-Myers Squibb, and Sanofi, research funding from ECTRIMS, and travel funding from Janssen.
DP is a member of the advisory board for Cognition and MS for Novartis and has received speaking honoraria from Biogen, Novartis, MedAhead and Bristol-Myers Squibb.
CE has received funding for traveling and speaker honoraria from Biogen Idec, Bayer Schering Pharma, Merck Serono, Novartis, Genzyme and Teva Pharmaceutical Industries Ltd./sanofi-aventis, Shire; received research support from Merck Serono, Biogen Idec, and Teva Pharmaceutical Industries Ltd./sanofi-aventis; and serves on scientific advisory boards for Bayer Schering Pharma, Biogen Idec, Merck Serono, Novartis, Genzyme, Roche, and Teva Pharmaceutical Industries Ltd./sanofi- Aventis.
TU received financial support for conference travel and honoraria from Biogen, Novartis, Roche, Bristol Myers Squibb, and Merck, as well as support for research activities from Biogen and Sanofi. He also received support by the Czech Ministry of Health, the institutional support of a hospital research project (MH CZ-DRO-VFN64165), the Czech Ministry of Health project (NU22-04-00193) and the Charles University Cooperation Program in Neuroscience.
MV received speaker honoraria and consultant fees from Biogen, Novartis, Roche, Sanofi, and Teva, as well as support for research activities from Biogen. She also received support by the Czech Ministry of Health, the institutional support of a hospital research project (MH CZ-DRO-VFN64165), the Czech Ministry of Health project (NU22-04-00193) and the Charles University Cooperation Program in Neuroscience.
JK received research grants for multicentre investigator-initiated trials DOT-MS trial, ClinicalTrials. gov Identifier: NCT04260711 (ZonMW) and BLOOMS trial (ZonMW and Treatmeds), ClinicalTrials. gov Identifier: NCT05296161); received consulting fees for F. Hoffmann-La Roche, Biogen, Teva, Merck, Novartis and Sanofi/Genzyme (all payments to institution); reports speaker relationships with F. Hoffmann-La Roche, Biogen, Immunic, Teva, Merck, Novartis and Sanofi/Genzyme (all payments to institution); adjudication committee of MS clinical trial of Immunic (payments to institution only).
EMM serves on the editorial board of Neurology and Frontiers in Neurology and received research support from Stichting MS Research and ZonMW. Received speaker fees from Merck and Novartis.
HV has received research support from Merck, Novartis, Pfizer, and Teva, consulting fees from Merck, and speaker honoraria from Novartis; all funds were paid to his institution.
FB serves on the steering committee and is iDMC member for Biogen, Merck, Roche, EISAI, acts as a consultant for Roche, Biogen, Merck, IXICO, Jansen, Combinostics, has research agreements with Novartis, Merck, Biogen, GE, Roche and is co-founder and shareholder of Queen Square Analytics LTD.
MMS serves on the editorial board of Neurology, Multiple Sclerosis Journal and Frontiers in Neurology, receives research support from the Dutch MS Research Foundation, Eurostars-EUREKA, ARSEP, Amsterdam Neuroscience and ZonMW (Vidi grant, project number 09150172010056) and has served as a consultant for or received research support from Atara Biotherapeutics, Biogen, Celgene/Bristol Meyers Squibb, EIP, Sanofi, MedDay and Merck.
GP has received research grants from ECTRIMS, MAGNIMS, ESNR, and ZonMW.
All other authors declare they have no competing interests.

### Author Declarations

Ethics commitee of Amsterdam UMC gave ethical approval for this work

